# Modelling Vaccination Strategies for the Control of Marburg Virus Disease Outbreaks

**DOI:** 10.1101/2022.06.17.22276538

**Authors:** George Y Qian, W John Edmunds, Daniel G Bausch, Thibaut Jombart

## Abstract

Marburg virus disease is an acute haemorrhagic fever caused by Marburg virus. Marburg virus is zoonotic, maintained in nature in Egyptian fruit bats, with occasional spillover infections into humans and nonhuman primates. Although rare, sporadic cases and outbreaks occur in Africa, usually associated with exposure to bats in mines or caves, and sometimes with secondary human-to-human transmission. Outbreaks outside of Africa have also occurred due to importation of infected monkeys. Although all previous Marburg virus disease outbreaks have been brought under control without vaccination, there is nevertheless the potential for large outbreaks when implementation of public health measures is not possible or breaks down. Vaccines could thus be an important additional tool and development of several candidate vaccines is under way. We developed a branching process model of Marburg virus transmission and investigated the potential effects of several prophylactic and reactive vaccination strategies in settings driven primarily by multiple spillover events as well as human-to-human transmission. Our results show a low basic reproduction number which varied across outbreaks, from 0.5 [95% CI: 0.05 – 1.8] to 1.2 [95% CI: 1.0 – 1.9] but a high case fatality ratio. Of six vaccination strategies explored, a combination of ring and targeted vaccination of high-risk groups was generally most effective, with a probability of controlling potential outbreaks of 0.88 (95% CI: 0.85 - 0.91) compared with 0.65 (0.60 - 0.69) for no vaccination, especially if the outbreak is driven by zoonotic spillovers and the vaccination campaign initiated as soon as possible after onset of the first case.

**Author Summary:** Marburg virus disease is a rare but acute haemorrhagic fever caused by Marburg virus. We developed a branching process model of Marburg virus transmission and used this model to investigate potential prophylactic and reactive vaccination strategies in settings driven primarily by multiple spillover events as well as human-to-human transmission. We calculate a low basic reproduction number which varied across outbreaks, from 0.5 [95% CI: 0.05 – 1.8] to 1.2 [95% CI: 1.0 – 1.9].

Of the six vaccination strategies explored, a combination of ring and targeted vaccination of high-risk groups was generally most effective, with a probability of controlling potential outbreaks of 0.88 (95% CI: 0.85 - 0.91) compared with 0.65 (0.60 - 0.69) for no vaccination, especially if the outbreak is driven by zoonotic spillovers and the vaccination campaign initiated as soon as possible after onset of the first case.

## Introduction

Marburg virus disease (MVD) is an acute haemorrhagic fever caused by Marburg virus (genus *Marburg marburgvirus*, family *Filoviridae*), affecting humans and non-human primates (1–4). Marburg virus is zoonotic, maintained in nature in Egyptian fruit bats (*Rousettus aegyptiacus*), which are found across Africa (5). Although rare, sporadic cases and outbreaks occur, usually associated with exposure in mines or caves inhabited by colonies of Egyptian fruit bats (5–13). Secondary human-to-human transmission may occur through direct exposure to blood and body fluids or contaminated surfaces.

There have been 15 recognized MVD outbreaks to date, beginning in 1967 when infected green monkeys from Uganda were imported to Germany and Yugoslavia for harvesting of their tissues for vaccine production (3, 14) (Table A1). Excluding laboratory infections, the exposures of all index cases of outbreaks since then have occurred in Africa (5–13), with some outbreaks driven by recurring virus spillover from animals to humans and others primarily by human-to-human transmission. Figure 1 and Table A1 show the MVD case numbers observed during each of these outbreaks.

**Figure 1:**
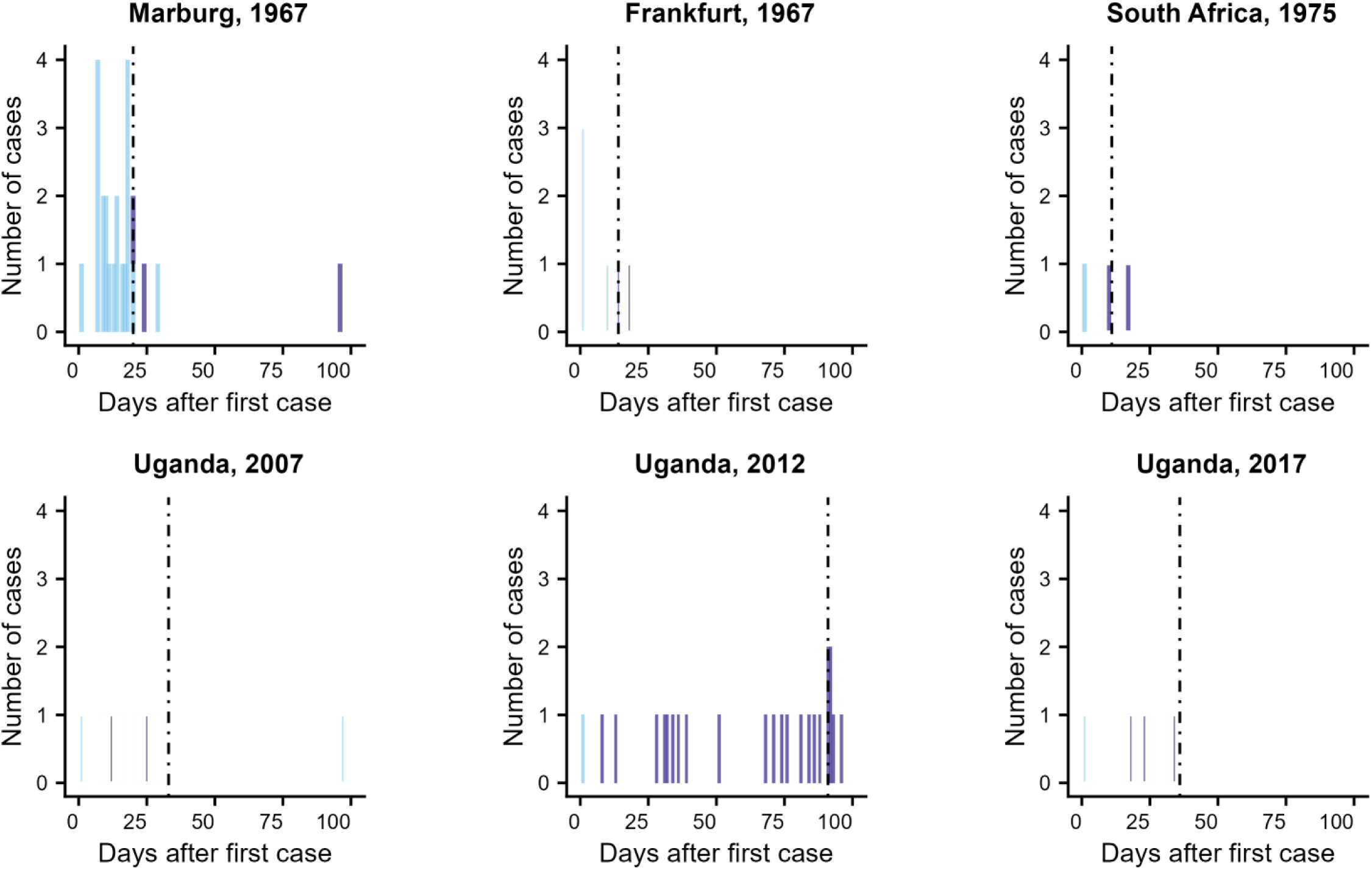
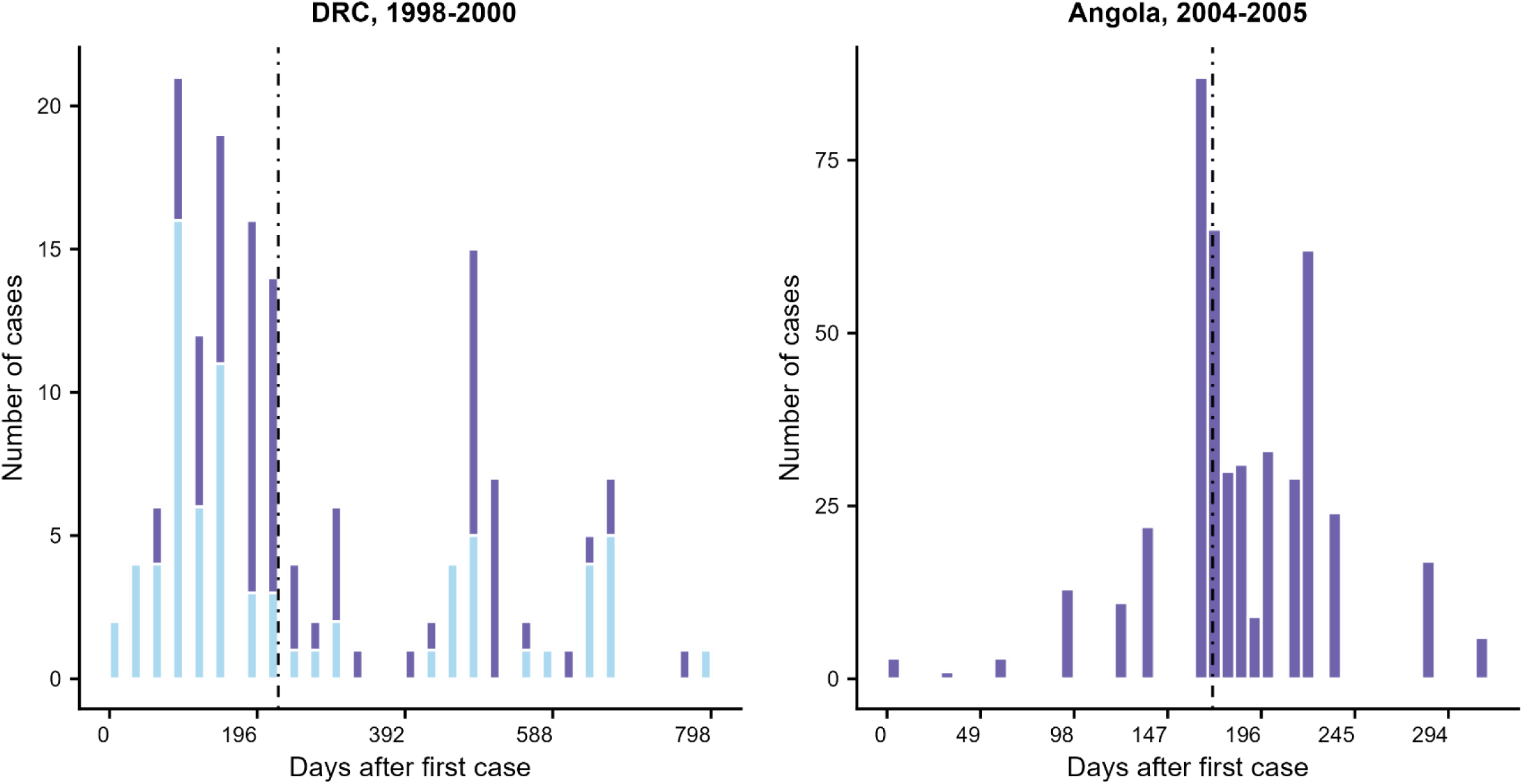
Incidence plots of Marburg virus disease outbreaks involving three or more confirmed or probable cases. Cases are plotted by onset date, except for the outbreak in Angola, which shows the day of reporting. Dashed lines represent the day on which interventions were put in place. Light blue bars indicate probable spillover infections to humans from animals and dark blue bars cases of human-to-human transmission.

No licensed vaccine for MVD currently exists, although several are under development (4). All previous outbreaks were controlled when transmission chains ended either naturally or through the introduction of public health and infection control measures (7, 15). Nevertheless, the 2004 outbreak in Angola, which registered 374 cases and 329 deaths (case fatality of 88 percent), illustrates the serious and explosive potential of Marburg virus. Furthermore, even in the smaller outbreaks, the high case fatality ratio could potentially be mitigated by vaccination (4).

The aims of this study were to estimate key epidemiological parameters of MVD, such as the reproduction number and serial interval, by collating data from all 15 previous human outbreaks and subsequently use this information to parameterise a model that is used to assess the impact of different vaccination strategies to control outbreaks.

## Methods

### Data

We used linelist data from all except one of the previous 15 outbreaks to estimate the serial interval of MVD. The one exception was the Angola outbreak that occurred between 2004-2005; here, because no linelist was available, we used case numbers reported periodically by the World Health Organisation (reported in (16), for instance).

From the linelist data, we identified discernable infector-infectee pairs, obtained the difference between the dates of infection of each pair and fit a gamma distribution to these differences. We also used the linelist data to obtain the number of zoonotic introductions seen in each outbreak and, together with knowledge of the duration of outbreak, calculated the rate of introductions. These data can be found in the following repository: https://github.com/GeorgeYQian/MVD-Branching-Process-Model-Repository

### Rate of zoonotic introductions

Since several outbreaks were driven primarily by zoonotic introductions, while others were largely caused by human-to-human transmission (Figure 1), we estimated rates of zoonotic introductions to represent both these types of outbreaks.

For outbreaks dominated by contact with animals, we used data from the 1998-2000 outbreak in the Democratic Republic of the Congo (DRC) (6), which was driven by miners being infected through spillovers from bats. It was reported that 27% of infected miners from this outbreak had contact with another infected individual (6), from which we infer that 73% of all infected miners were spillover cases. Given that the outbreak lasted 2 years (with the first case identified in October 1998 and the last in September 2000 (6)), we divided the number of spillover cases by this duration to estimate the rate of introductions during the DRC outbreak. We use this as a typical rate of introductions for spillover-driven outbreaks. Other outbreaks likely involved a single spillover event subsequently driven by human-to-human transmission, typified by the large 2004 outbreak in Angola. To obtain the rate of introductions, we therefore divided the number of spillover cases by the duration of these outbreaks.

### Time from first case to interventions

For each outbreak, we estimated the date on which interventions were put in place. During earlier outbreaks this was simply when the disease was acknowledged as being dangerous and highly transmissible, prompting changes in clinical, laboratory and infection prevention and control practices (14), or patients who showed symptoms consistent with other viral haemorrhagic fever and were treated accordingly, for instance through application of case isolation and barrier nursing (17). For later outbreaks we used the day on which response teams were deployed to the region of the outbreak as the intervention date (while recognizing that local control efforts were often already underway). We calculated the median time delay between onset of the first case and the beginning of interventions across all outbreaks. However, intervention during several outbreaks, including those in Angola (2004-2005) (16, 18), DRC (1998-2000) (6) and Uganda (2012) (19) took place several weeks after the median. Hence, as a sensitivity analysis, we took the 75th percentile of this delay to intervention and modelled this scheme.

### Factors affecting outbreak size

The number of cases in each MVD outbreak is presumed to be dependent on several factors, including the number of zoonotic introductions, delay from first case to intervention and calendar year in which the outbreak occurred. The impact of armed conflict was also noted as a possible factor for the two largest MVD outbreaks, in DRC and Angola (6,18,20). However, as conflict in both these regions had officially ended shortly before the outbreaks occurred, we speculate that recent armed conflict in the outbreak region may also have influenced the final outbreak size.

We used a negative binomial generalised linear model (GLM) to test if the total number of secondary cases (i.e. excluding zoonotic introductions) was linked with these covariates. A negative binomial distribution for the response variable was preferred to a Poisson because of the high over-dispersion observed in this variable. Note that such dispersion was not observed in daily case incidence within individual outbreaks, which we later-on modelled using a Poisson branching process.

### Branching process model

We used a branching process to model MVD transmission over time. New infections generated at any time, *t*, are governed by the force of infection *λt*, which is determined by previous case incidence *ys* (*s* = 1, …, *t*-1), the serial interval distribution (denoted by *w*, its probability mass function), and the reproduction numbers *Rs* as (21):

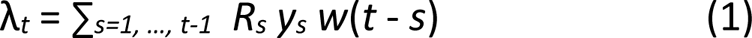

New secondary cases at time *t* are then drawn from a Poisson distribution so that:

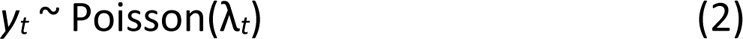

Equation (1) shows that the reproduction number *Rs* is allowed to vary over time. This is used to distinguish, in any given outbreak, two phases: a first one, during which transmission is maximum (*Rs* = *R*0, the basic reproduction number), and a second one during which intervention reduces transmission by a factor *E*, the intervention efficacy, so that:

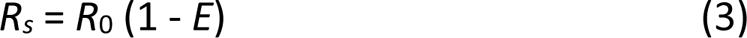

Intervention is defined, in this context, as the implementation of measures such as case isolation, contact tracing and barrier nursing. We assume that depletion of the susceptible population is negligibly low, given the low number of cases compared to the population of each affected community.

The date at which interventions started reducing transmission is outbreak dependent, and was obtained through investigation of the respective, publicly available outbreak reports (3,6–12,16,17,19,22,23). Thus, in our model, the reproduction number decreases from *R0* to *Rs* after this date.

The model also incorporates a constant rate of introductions *γ* in which newly introduced cases (presumed spill-over events) are also Poisson distributed.

Hence, the number of new cases at time *t*, *Y_t_*, is the sum of the primary and secondary cases:

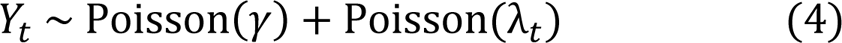

### Parameter estimation

We used an Approximate Bayesian Computational (ABC) framework for estimating the basic reproduction number, *R0*, and *E* for each outbreak separately. To do this, we first determined the delay to implementation of interventions, the duration of the outbreak and rate of introductions for each outbreak. The priors used were: *R0* ∈ U(0,3) and *E* ∈ U(0,1). The summary statistic used was the absolute difference between the total number of cases observed in a simulation and the actual number reported during the outbreak. Parameter values were retained as part of the posterior sample if this difference was within 10% of the actual value:

|*n_simulated_*−*n_outbreak_*| < 0.1 *n_outbreak_*. 5000 posterior samples were retained in this way per outbreak.

Along with the reproduction numbers, we also estimated the dispersion parameter, *k*, a measure of the variation in the number of susceptibles infected by each infected individual across an outbreak, as well as the serial interval. For *k*, we identified 18 chains of transmission from the outbreak in DRC (1998-2000) (6).

We determined the number of secondary cases generated by each index case and fit a negative binomial distribution to these (see (24)). The size parameter of this distribution represents the dispersion parameter, *k* (24). For the serial interval, we identified 26 infector-infectee pairs from linelist data obtained from MVD outbreaks in DRC, Kenya, South Africa, Belgrade and Uganda (2012) and fit a gamma distribution to the time period between the dates of onset for these infector-infectee pairs (25).

We then modified the model to include the effects of vaccination on transmission. Vaccination reduces the R value associated with each case by the vaccine efficacy (*VE*) corresponding to that case, on top of any reduction due to intervention efficacy (*E*). Six vaccination schemes were simulated:

1. *Prophylactic targeted vaccination of high-risk groups*. This scheme involves the vaccination of healthcare workers as well as individuals who reside near or work in mines or caves prior to the beginning or an outbreak. We estimate that, across all MVD outbreaks, approximately 6% of cases were healthcare workers and 12% were individuals living near or working in mines or caves (this excludes the outbreak in Angola due to lack of data).
2. *Prophylactic mass vaccination*. The second scheme was prophylactic mass vaccination of the entire community prior to the outbreak.
3. *Ring vaccination*. This is the first of the reactive vaccination strategies that we simulate. A proportion of contacts are vaccinated after the date of intervention. This proportion depends on how extensively case reporting is carried out, as well as on vaccination coverage.
4. *Reactive targeted vaccination*. This scheme entails vaccination of the same high-risk groups as in scheme #1 above, but done reactively after an MVD outbreak has begun. In our model, vaccination is simulated only after the date of intervention.
5. *Reactive mass vaccination*. This is mass vaccination simulated only after intervention has begun.
6. *A combination of ring and reactive targeted vaccination schemes*. We assumed that no waning of immunity occurred after vaccination, nor depletion of susceptibles.

The vaccination parameters in our model were:

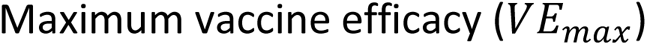

*VE_max_* of a vesicular stomatitis virus (VSV)-based vaccine expressing the MARV glycoprotein (VSV-MARV) was found to be 100% in non-human primates (NHPs) (2). As this is unlikely to be observed in the field, we adjusted downward to a *VE_max_* of 90% in the base case.

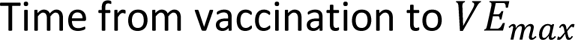

The time from vaccination, when efficacy is 0%, to maximum efficacy was 7 days, from Phase I trials on NHPs of the MVD vaccine (2). This is reflected in our logistic curve representing the vaccination efficacy over time (Technical Appendix Figure A1).

### Vaccine coverage

The level of coverage that is likely to be achieved in an outbreak setting is difficult to estimate. In two reviews of oral cholera vaccine, uptake coverages ranged between 61%-100% for at least 1 dose and 46%-92% for two dose coverage in outbreak settings. Hence in this analysis, in the base-case we assume that targeted and ring vaccination coverages would be 70%, at the lower end of the one-dose coverage, since MVD vaccines currently in the pipeline are injectable, rather than oral (as is the case with oral cholera vaccines).

Given logistical difficulties and possible vaccine shortages (26), we would expect a lower rate of coverage (assumed to be 50%) in the case of a mass vaccination strategy (both reactive and prophylactic). We increased or decreased these coverages by 20% in the sensitivity analyses.

### Time between vaccination and infection

The time between vaccination and infection is not independent in a reactive programme as contacts of cases (in the case of ring vaccination) or the wider community (mass vaccination) are vaccinated in response to a case. Thus many people will be vaccinated around the time that they are infected, lowering vaccine efficacy. We have taken this into account by assuming that, amongst individuals who become infected, there was an average delay between vaccination and infection of 20 days (s.d. 5 days) for prophylactic and 9 days (s.d. 4 days) for reactive strategies. These distributions were chosen so that vaccine efficacy varied during reactive vaccination, with most of the efficacy values lying on the slope of the logistic curve fit of vaccine efficacy (see next section), while for prophylactic vaccination, over 99% of values were within 0.5% of the maximum vaccine efficacy.

Data from a previous ring vaccination trial for an Ebola virus disease vaccine in Guinea showed an average delay from vaccination to subsequent infection (in those that got infected) in the rings of 5.7 days (s.d. 5.0 days) (27). We also examined this delay distribution as a sensitivity analysis.

### Logistic curve fit of vaccine efficacy

Timing is crucial for the success of vaccination strategies, particularly those that are reactive. Hence, we modelled the vaccine efficacy from vaccination to infection using a logistic curve of the form (Figure A1 in the Technical Appendix):

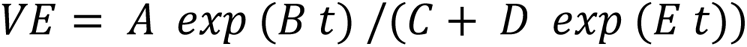

where *VE* is the vaccine efficacy as a function of time, *t*, in days and *A, B, C, D* and *E* are constants to be determined through a fitting procedure. A self-starting nonlinear least squares approach (specifically, the Levenberg-Marquardt algorithm) was used to fit this curve so that *VE* increases from 0 to the within 0.5% of the maximum *VE*, denoted *VE_max_*, in 12 days (see (2)) and subsequently tends towards this maximum.

In our model, each infected and vaccinated individual is assigned a vaccine efficacy, sampled from a particular region of this logistic curve. Which portion of this curve is sampled depends on the vaccination strategy employed. In general, prophylactic strategies result in higher vaccine efficiencies than reactive vaccination, since vaccines are administered before the beginning of an outbreak and so efficacy is assumed to have reached a maximum. We therefore took samples of *VE* from the top of the logistic curve (Technical Appendix). On the other hand, individuals vaccinated reactively in response to an outbreak are more likely to be infected before peak *VE* has been reached (here, we assume this period to be 12 days, based on (2)). Hence, we took samples of *VE* from a larger portion of the logistic curve (mainly its slope) in these scenarios (see Technical Appendix for further details).

### Forward simulations

We subsequently performed forward simulations to show the effects of these different vaccination schemes on potential outbreaks under both low and high introduction rates. For each forward simulation, we selected at random one set of parameters from the posterior distribution of one randomly selected MVD outbreak. We ran 5000 simulations per vaccination scheme in this manner.

We compared the distributions of simulated case numbers after implementing the six vaccination strategies described above with the no-vaccination scheme. We also estimated the proportion of controlled outbreaks predicted under each scheme. We considered an outbreak to be controlled if, after simulating for 365 days, the force of infection (see Equation 1) was less than 0.05, considered negligibly low.

The code is available on https://github.com/GeorgeYQian/MVD-Branching-Process-Model-Repository

## Results

### Epidemiological parameters

The estimated rate of zoonotic introductions was 0.06 per day and 0.003 per day during the DRC and Angola outbreaks, respectively. Across all outbreaks, the median delay between onset of the first MVD case and beginning of interventions was 21 days.

The fitted gamma distribution of the serial intervals had a mean of 9.2 days and standard deviation of 4.4 days (Figure A2, Technical Appendix). The median value of *R*_0_ was 0.8 [95% CI: 0·08–1·8] (Figure 2), while the median value of *R_s_*, the post-intervention reproduction number, was 0.3 [95% CI: 0·01–1·3], from Equation 3. However, these estimates have little practical value due to the large heterogeneity across outbreaks.

**Figure 2:**
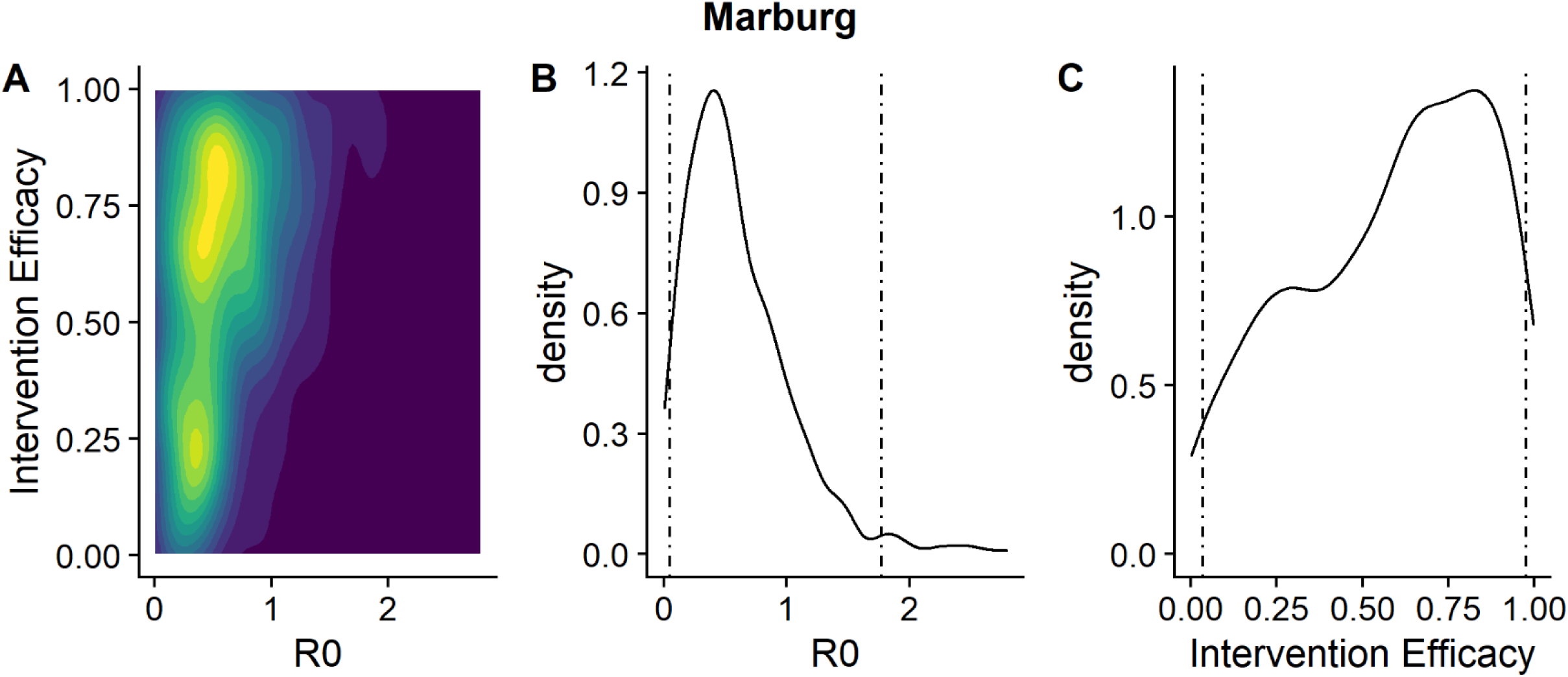
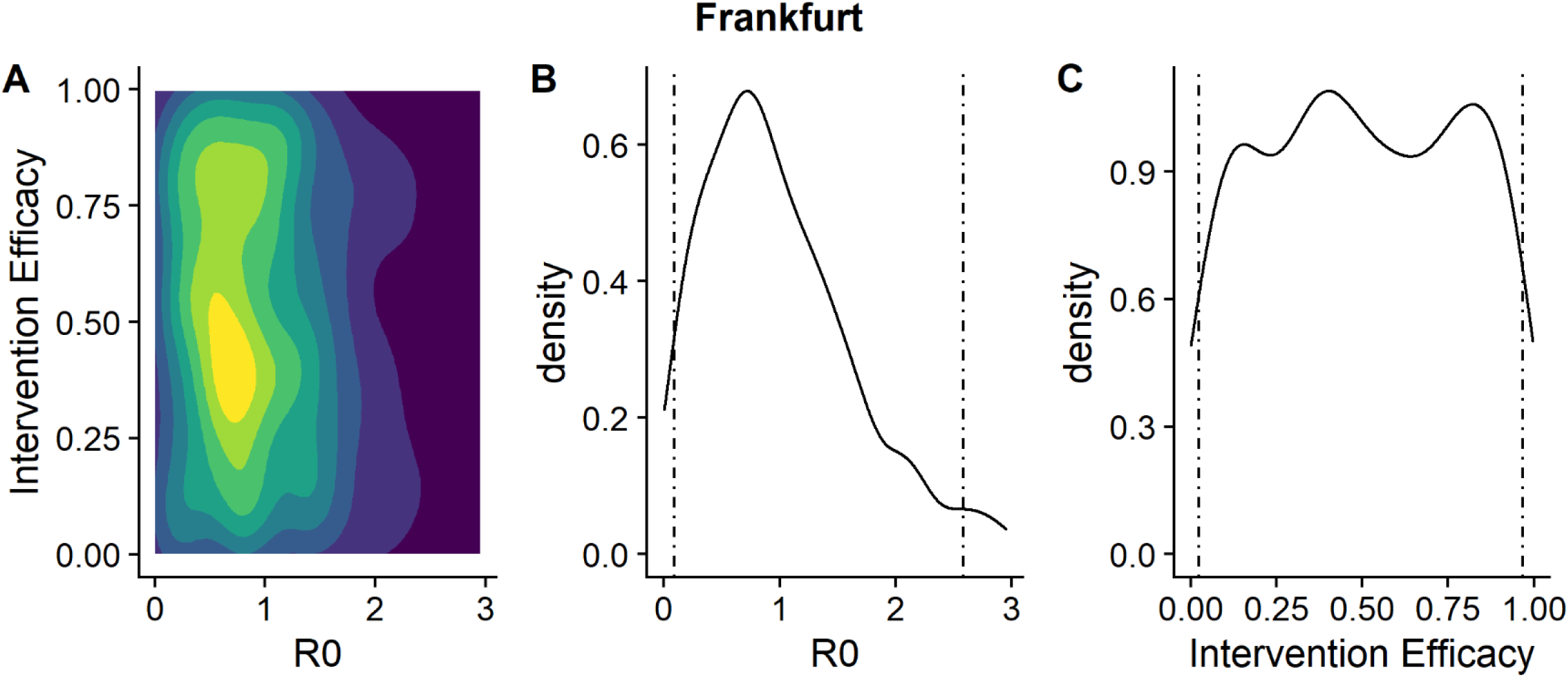
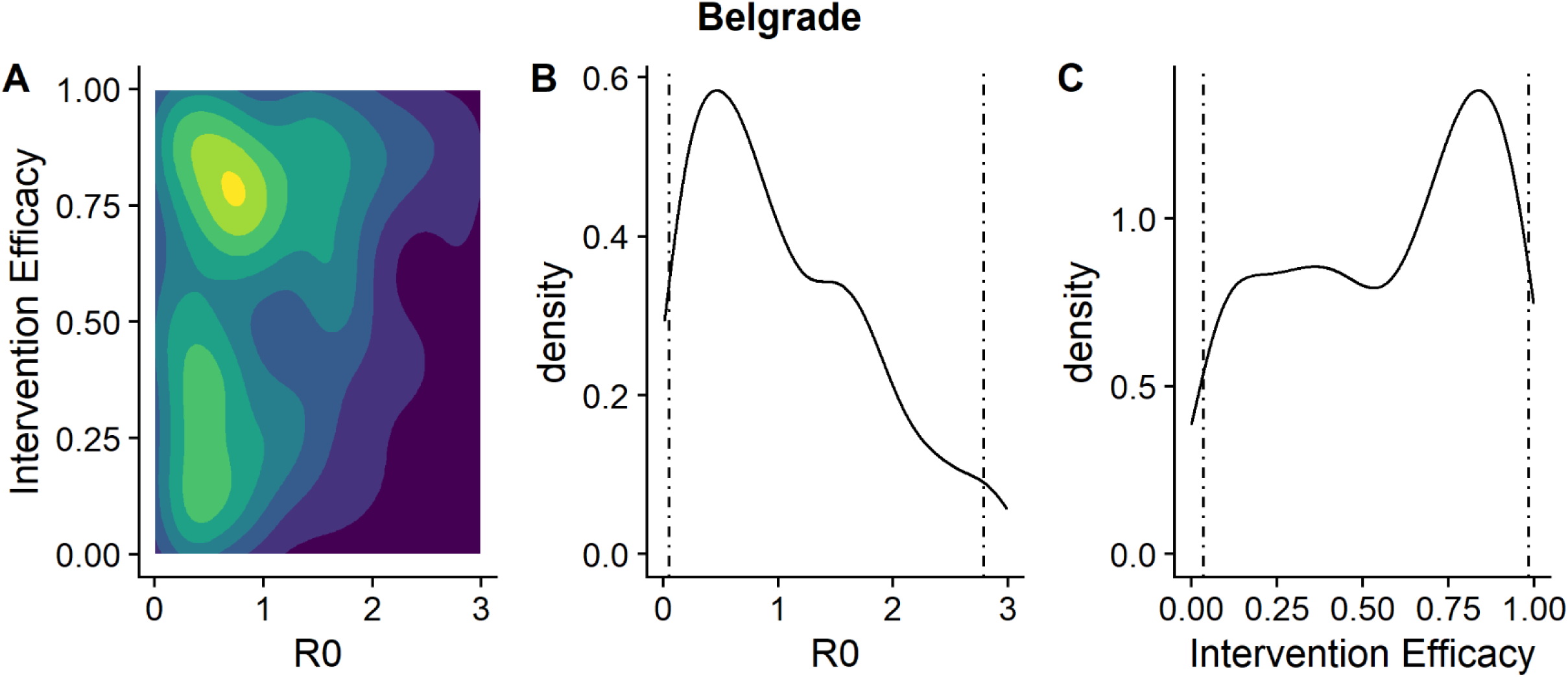
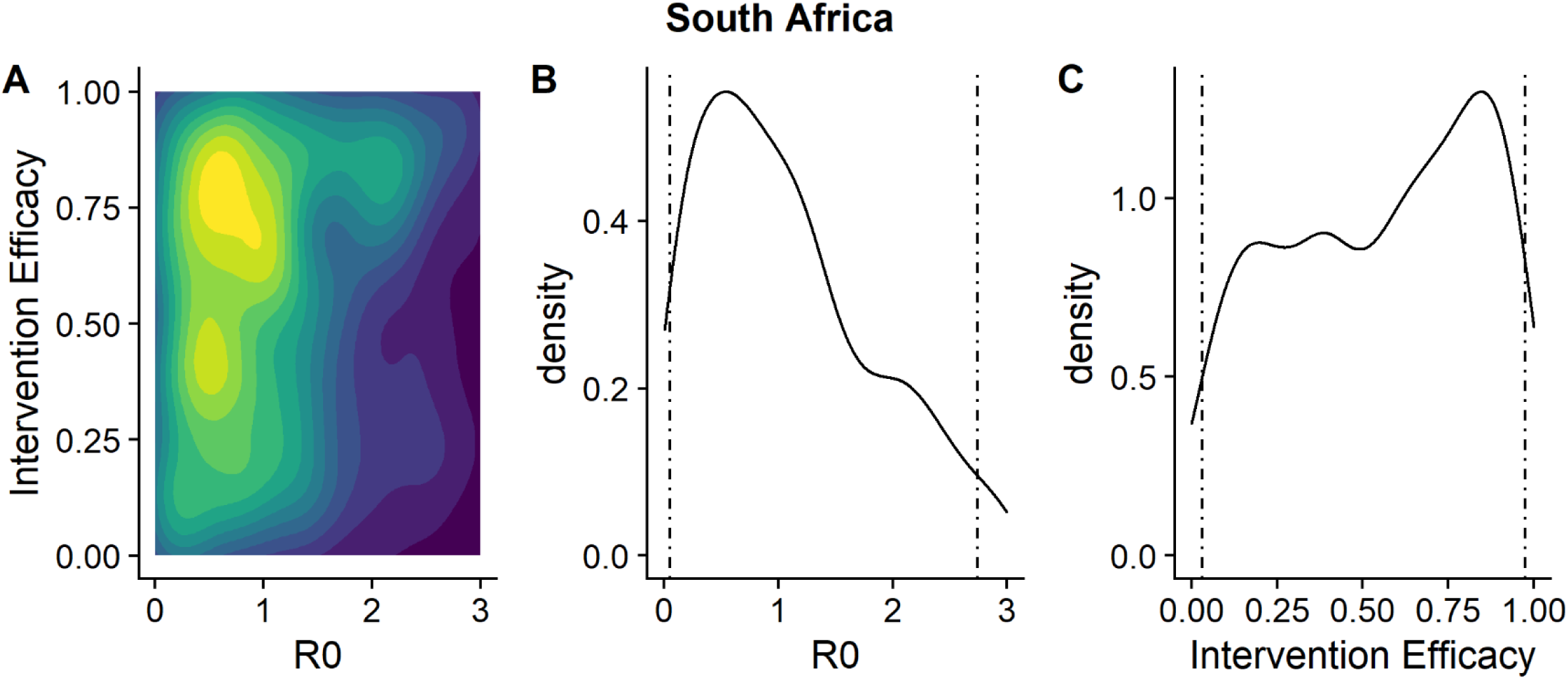
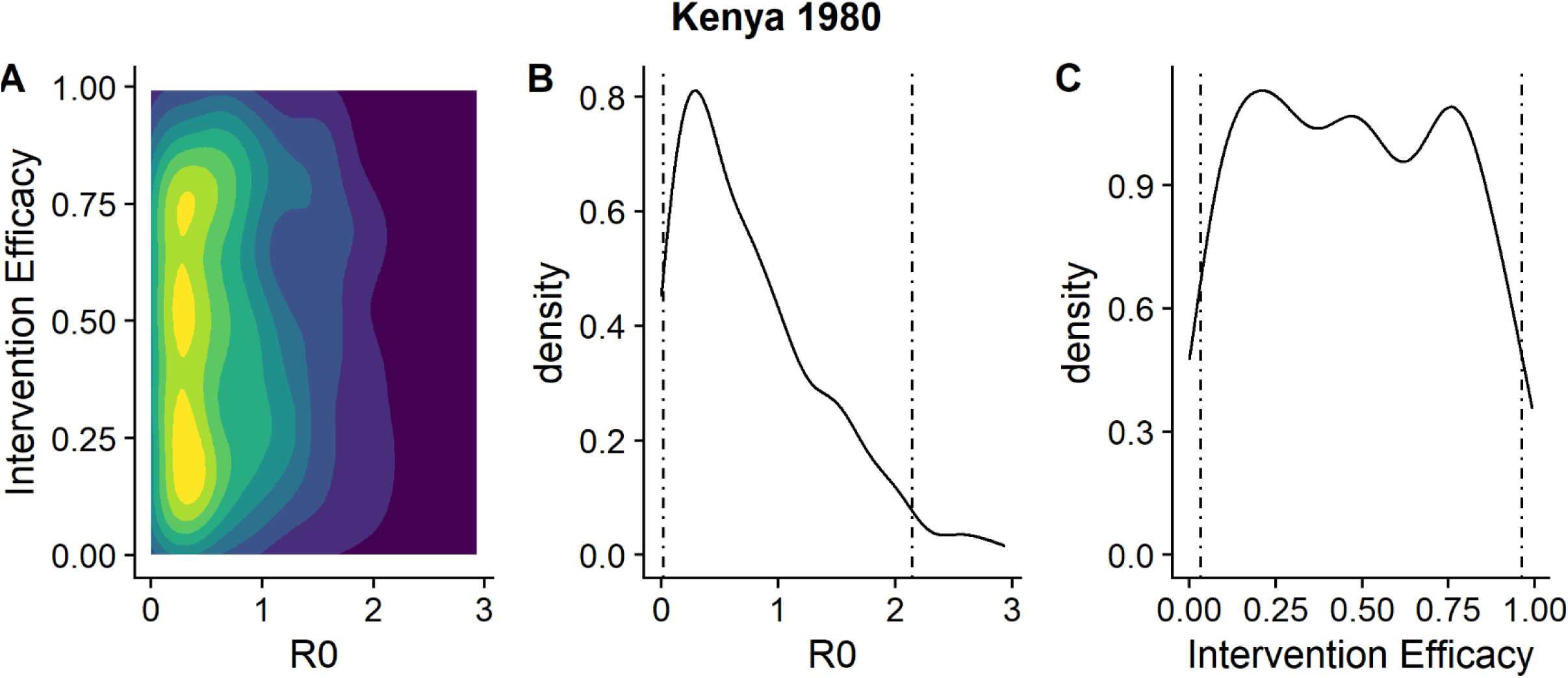
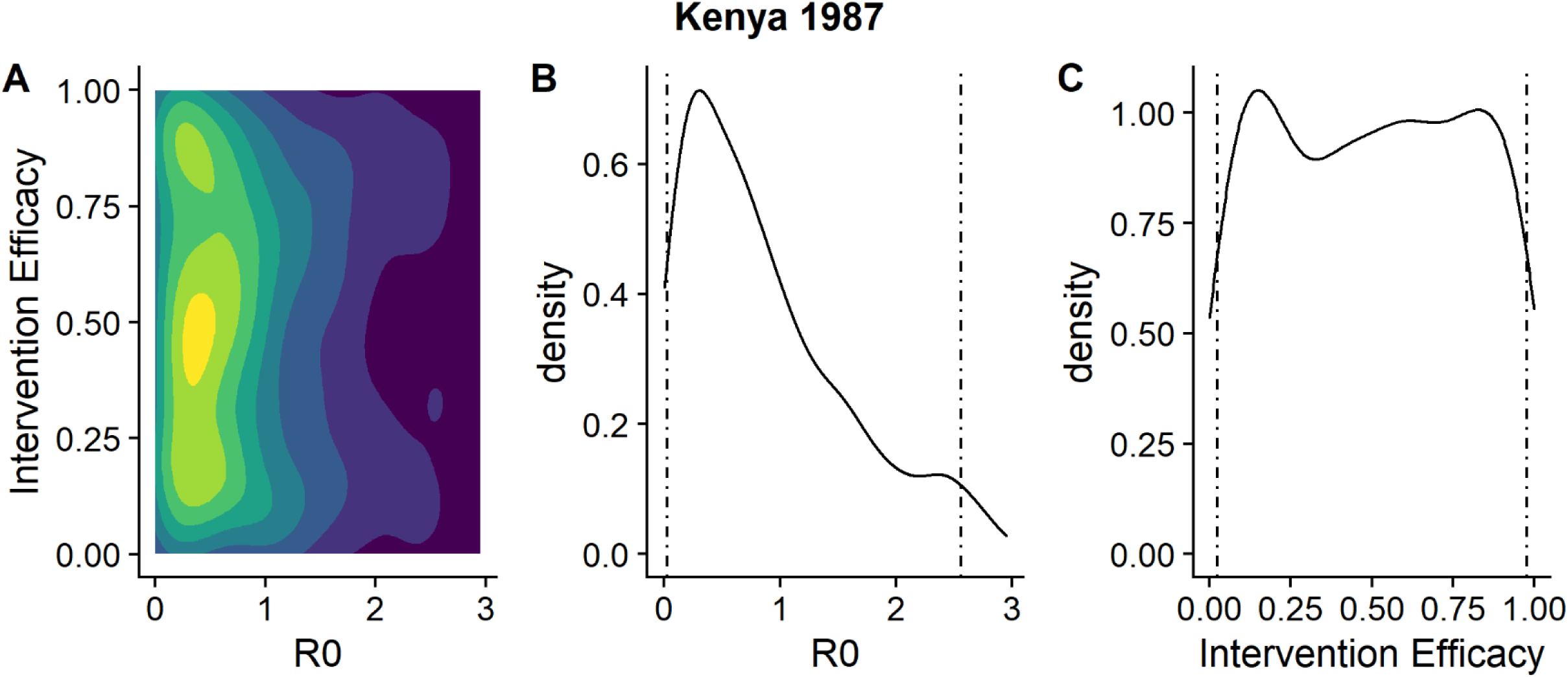
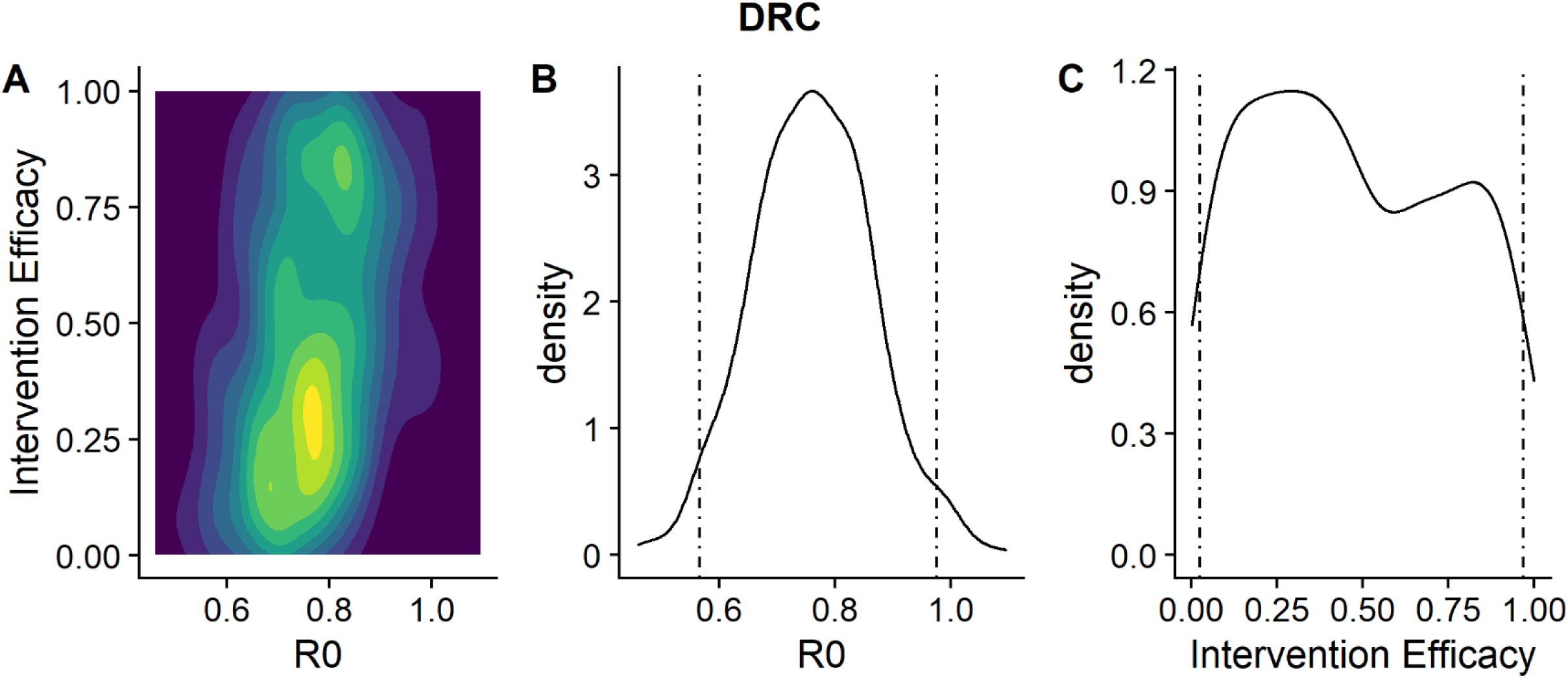
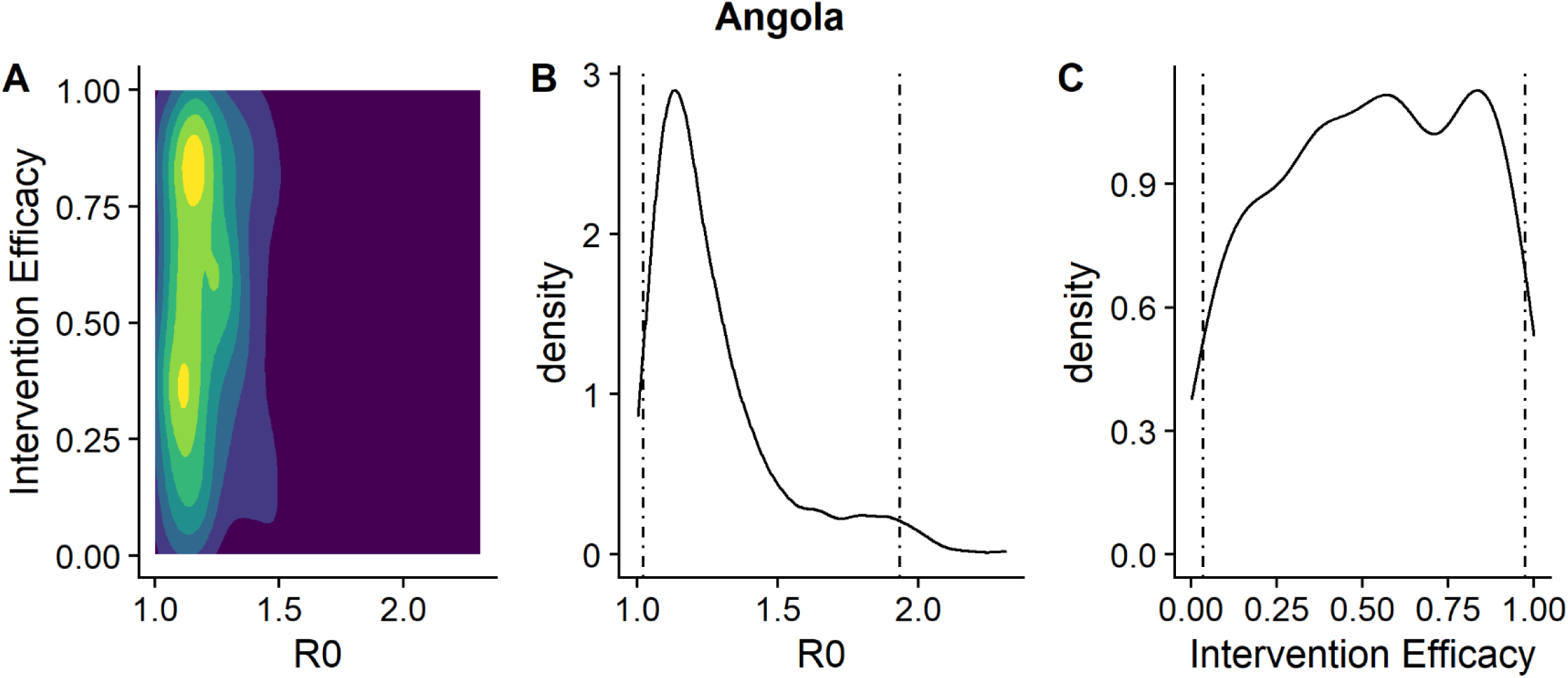
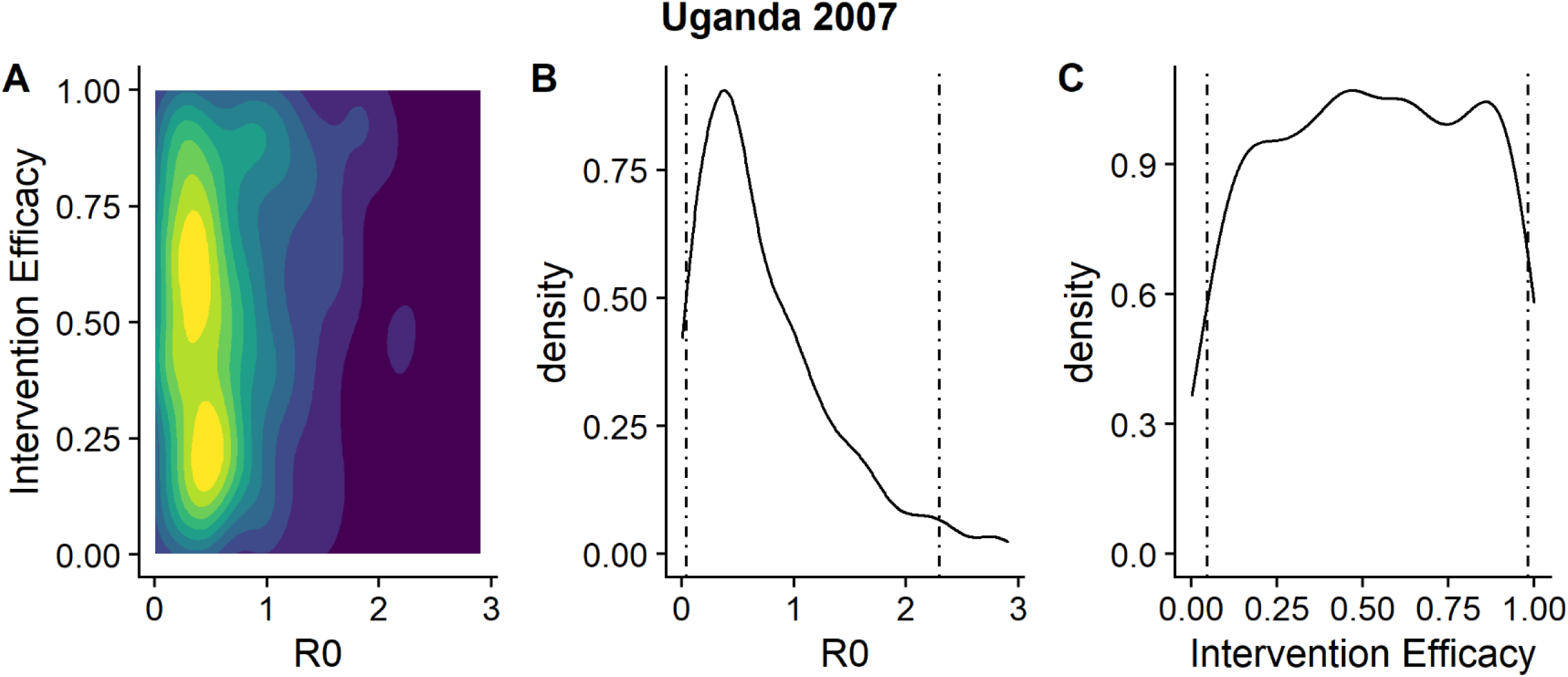
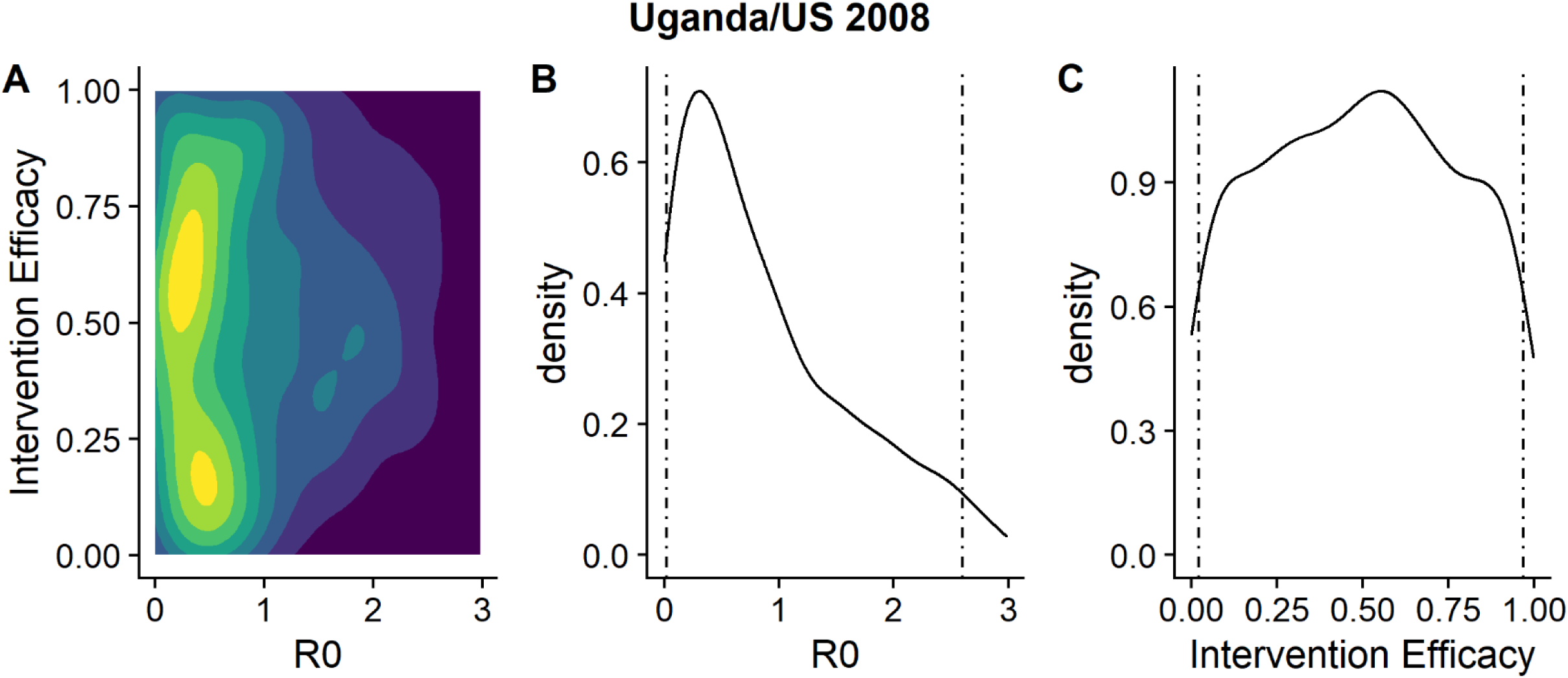
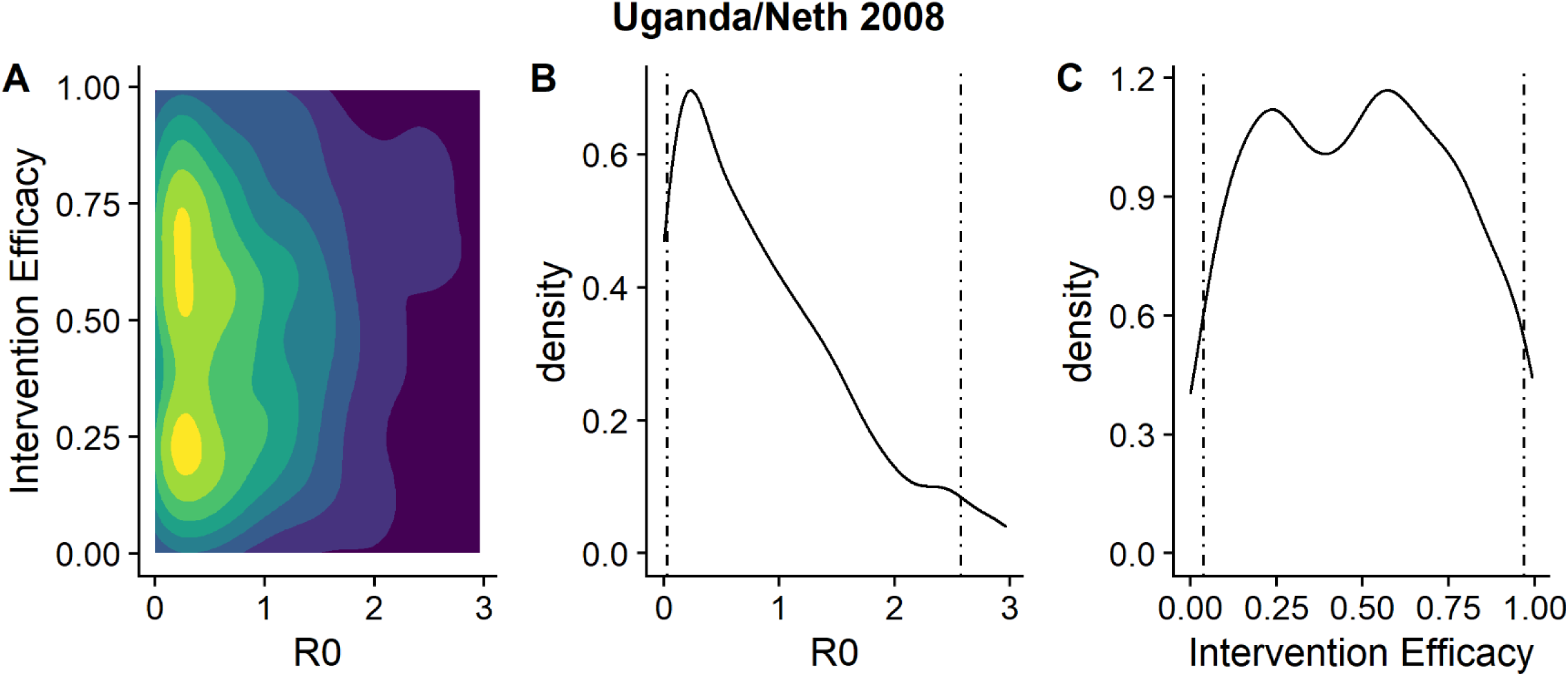
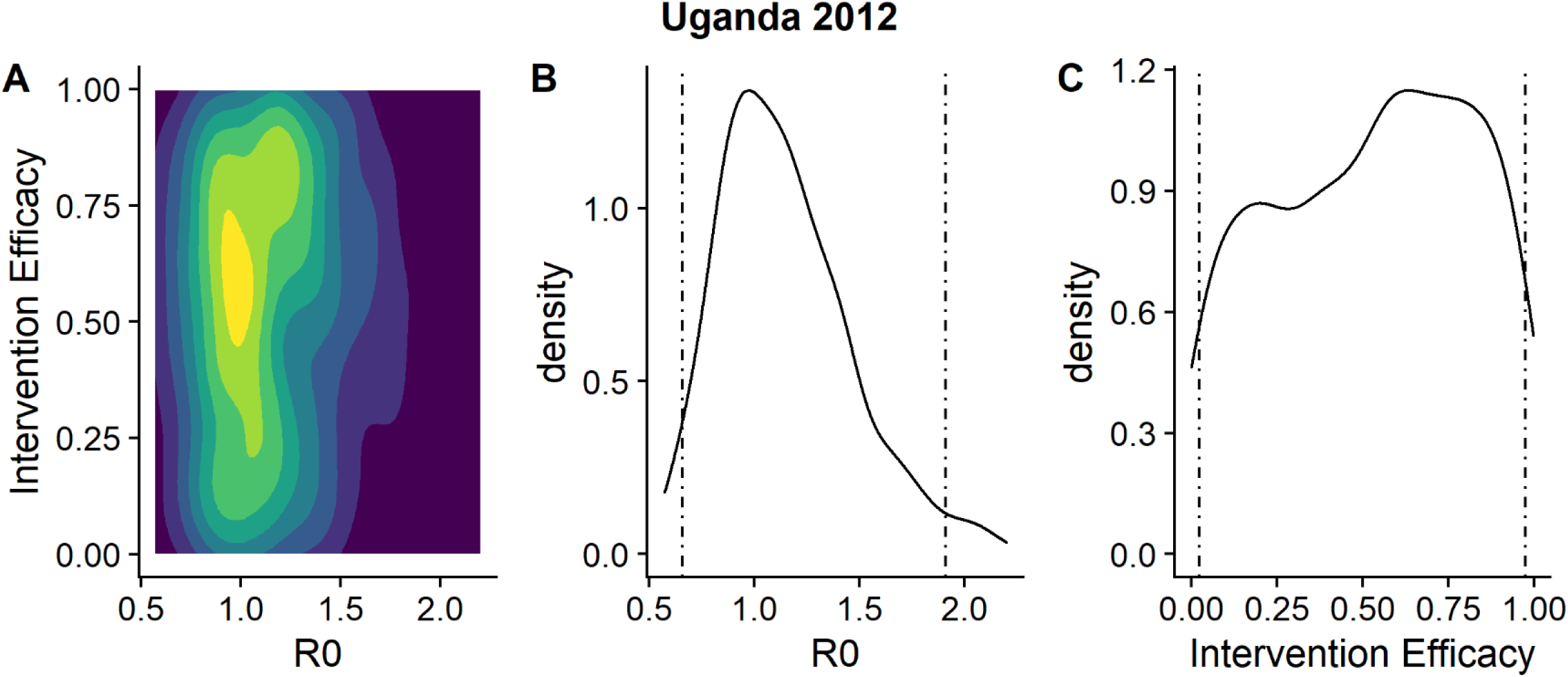
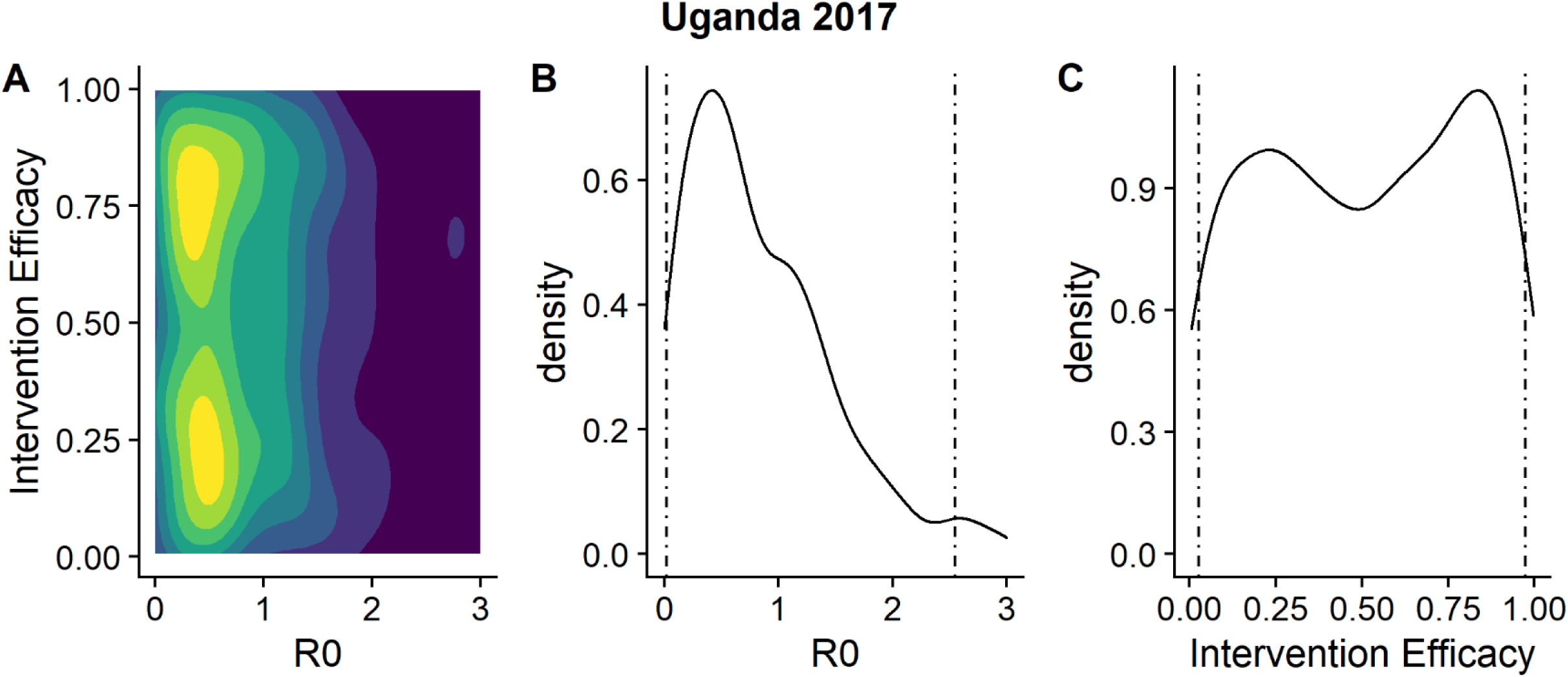
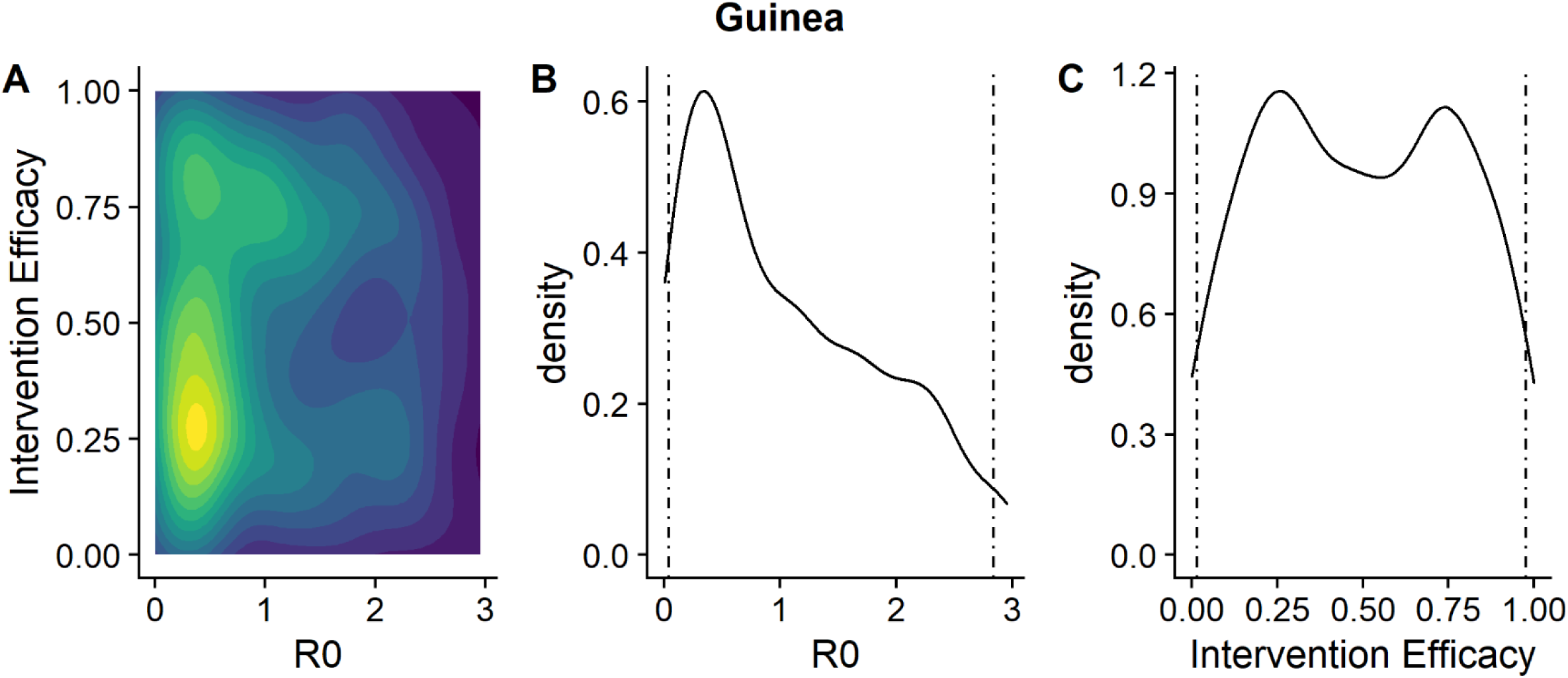
A. Posterior distribution of the basic reproduction number and intervention efficacy combined, B. marginal posterior distribution of the basic reproduction number and C. marginal posterior distribution of the intervention efficacy for each outbreak, in chronological order. Dashed lines indicate 95% confidence intervals.

Instead, it is more informative to inspect the distributions of *R_0_* and the post-intervention reproduction number for each previous outbreak: these are shown in Figure 2 and summarised in Table A2 (Technical Appendix). The value of *R*_0_ for each individual outbreak ranged from 0.5 [95% CI: 0.05 – 1.8] to 1.2 [95% CI: 1.0 – 1.9], while the post-intervention reproduction number ranged from 0.2 [95% CI: 0.006 – 0.7] to 0.6 [95% CI: 0.03 – 1.5]. For the majority of outbreaks, the median value of *R_0_* was less than 1, indicating there was little person-to-person transmission even before interventions. Consequently, the intervention efficacy is difficult to estimate during these outbreaks. There were, however, two outbreaks where *R_0_* was likely to have been greater than 1: namely the Angolan outbreak in 2004-5 and Ugandan outbreak in 2012, where *R_0_* was 1.2 (95% CI: 1.0-1.9) and 1.1 (95% CI: 0.7-1.9) respectively. For these outbreaks, our estimates of the intervention efficacy were skewed towards higher values (Figure 2). Finally, we estimated the value of *k* to be between 0.52 and 0.67 (Technical Appendix). These parameters are summarised in Table 1.

**Table 1:** Epidemiological parameters associated with Marburg virus and Ebola virus diseases

| Parameter | Marburg virus disease | Ebola virus disease |
| --- | --- | --- |
| **Case fatality ratio** | 53.8% [95% CI: 26.5 - 80%]<br>(32) | 65.0% [95% CI: (54.0–76.0%)]<br>(All Outbreaks)<br>(32) |
| **Serial Interval**<br>**(Gamma distribution)** | Mean: 9.2 days<br>Standard Deviation: 4.4 days | Mean: 11.6 days<br>Standard Deviation: 6.3 days*<br>(33) |
| **Incubation Period (Range of**<br>**central values, (range))** | 5-10 (2- 21) days (15,31) | 5.3–12.7 (1–21) days*<br>(34) |
| **Basic reproduction number** | 1.2 [95% CI: 1.0 – 1.9]<br>(Angola, 2004-5)<br>0.76 [95% CI: 0.57-0.98]<br>(DRC, 1998-2000)<br>1.1 [95% CI: 0.66 – 1.9]<br>(Uganda, 2012)<br>Median estimates range<br>from 0.51 to 1.2, depending<br>on outbreak | 1.71 [95% CI: 1.44, 2.01]<br>(Guinea)<br>1.83 [95% CI: 1.72, 1.94]<br>(Liberia)<br>2.02 [95% CI: 1.79, 2.26]<br>(Sierra Leone)* (33) |
| **Dispersion parameter** | 0.52 - 0.67 | 0.18 (Guinea) (24) |
| **Maximum vaccine efficacy** | 100% (on Nonhuman Primates) (2) | 100% (27) |
| **Days between vaccination and maximum efficacy** | 7 (2) | 10 (27) |

**\*Ebola virus disease outbreak in West Africa, beginning in 2013**

### Factors influencing outbreak size

The negative binomial regression suggested that, of all factors that we investigated, the delay to intervention was the only statistically significant factor influencing the size of MVD outbreaks (p < 0.001). Specifically, an increase of one day to this delay resulted in the outbreak size increasing by a factor of 1.03 (the incidence rate ratio). The pseudo-*R*^2^ value for this regression was 0.74. These results are shown in Table 2.

**Table 2:** Values of coefficients for the negative binomial regression (with a natural log link), for a model with the following covariates: number of zoonotic introductions, delay to interventions, calendar year of outbreak and whether armed conflict occurred shortly before. The null deviance was 57, residual deviance 15 and hence the pseudo-*R*^2^ value was 0.74.

| **Covariates** | **Estimated value** | **Standard error** | **Lower CI (2.5%)** | **Upper CI (97.5%)** | **p-value** |
| --- | --- | --- | --- | --- | --- |
| **Model intercept** | 41 | 44 | -46 | 130 | 0.36 |
| **Number of introductions** | -0.036 | 0.033 | -0.10 | 0.029 | 0.27 |
| **Delay to interventions** | 0.028 | 0.0080 | 0.012 | 0.043 | <0.001 |
| **Calendar year of outbreak** | -0.020 | 0.022 | -0.064 | 0.023 | 0.36 |
| **Armed conflict** | 1.3 | 1.5 | -1.7 | 4.3 | 0.41 |

### Simulations of vaccination strategies

The proportion of controlled outbreaks in the absence of any vaccination strategy was 0.91 (95% CI: 0.88 - 0.93) and 0.65 (CI: 0.60 - 0.69) when simulating low and high rates of introductions, respectively. Most vaccination strategies resulted in a significant increase in this proportion, in particular the combined ring and targeted strategy, with values of 0.99 (CI: 0.97 - 0.99) and 0.88 (CI: 0.85 - 0.91), and the prophylactic mass strategy, with values of 0.99 (CI: 0.97 - 0.99) and 0.83 (CI: 0.79 - 0.86), for low and high rates of introductions, respectively (Figure 3a).

**Figure 3a:**
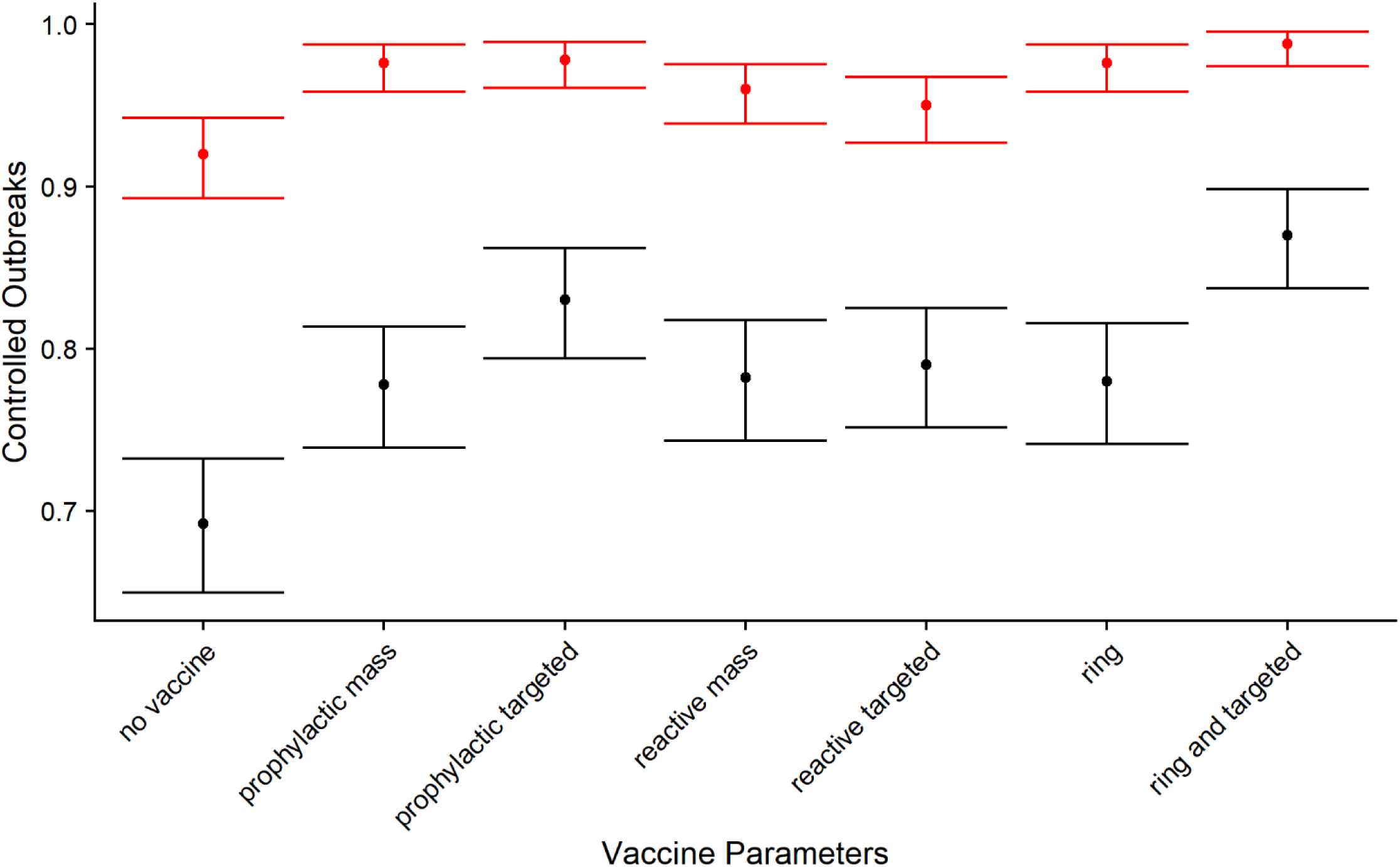
Proportion of controlled outbreaks predicted under different vaccination strategies, when the rate of zoonotic introductions is low (red bars) and high (black bars).

The median number of cases in the absence of any vaccination strategy was 3 (95% CI: 3-4) and 37 (CI: 34 - 40) for low and high rates of introductions, respectively. Under the low rate of introductions scenario, there was a small but significant decrease in this median to 2 cases for the prophylactic mass and targeted vaccination strategies, while there was also a small but significant decrease observed for all vaccination strategies when simulating a high rate of introductions, with the exception of the ring, reactive mass and reactive targeted vaccination schemes. In particular, the median number of cases under the prophylactic targeted strategy was 28 (CI: 27-29) (Figure 3b).

**Figure 3b:**
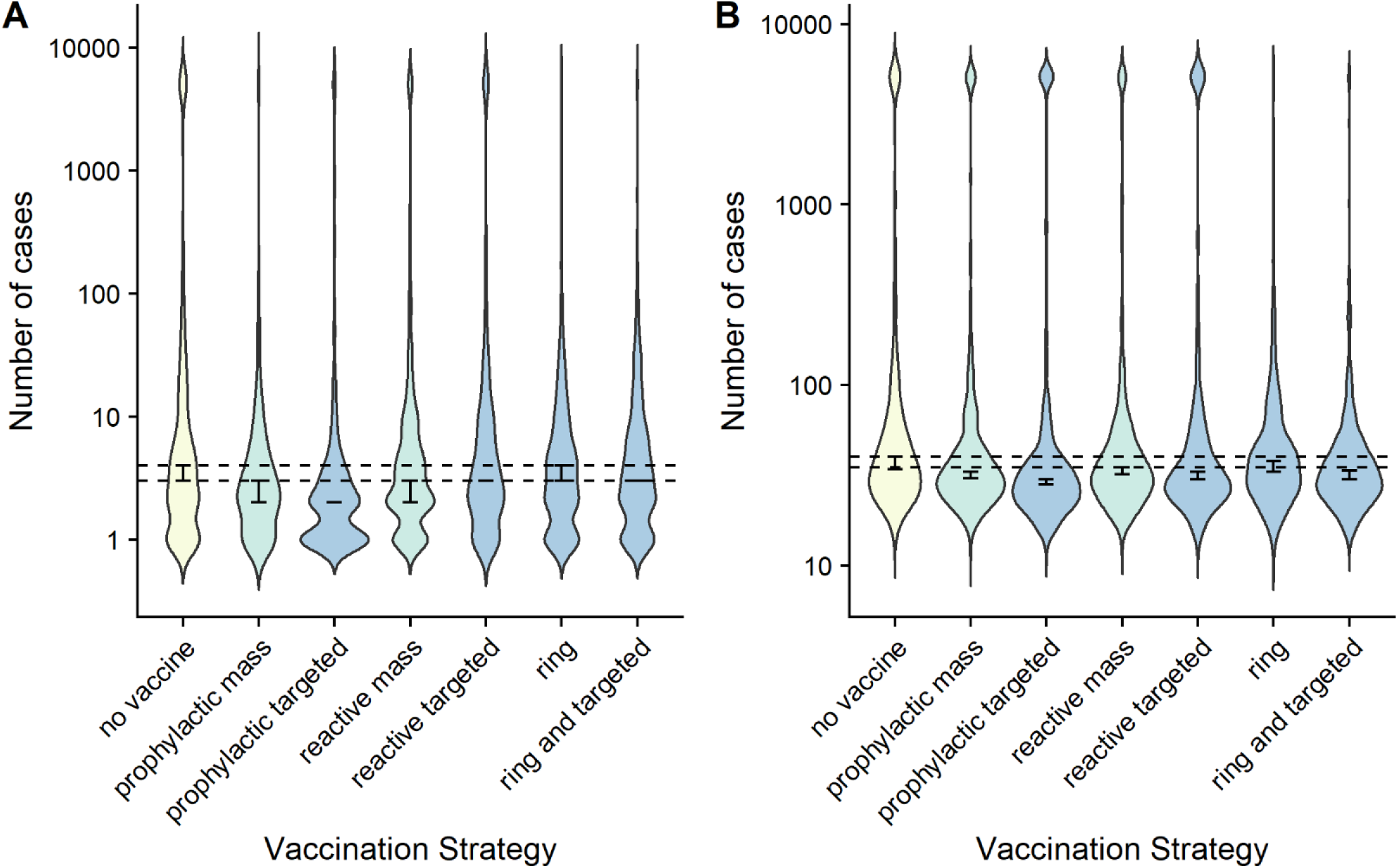
Proportion of controlled outbreaks predicted under different vaccination strategies, when the rate of zoonotic introductions is low (Panel A) and high (Panel B). The dashed lines represent the upper and lower bounds of the 95% confidence intervals in the absence of vaccination (“no vaccine”).

#### Sensitivity analyses

Varying the vaccination parameters had the following effects:

Reducing the delay between vaccination and infection to 5.73 days (s.d. 5.03 days) did not result in any significant differences from baseline values in either the proportion of controlled outbreaks or the median number of cases across all simulated outbreaks (Tables 3 and 4).

**Table 3:** Proportion of controlled outbreaks for different vaccination schemes

| **Scheme** | **No vaccination** | **Prophylactic mass** | **Prophylactic targeted** | **Reactive mass** | **Reactive targeted** | **Ring** | **Ring and reactive targeted** |
| --- | --- | --- | --- | --- | --- | --- | --- |
| **Baseline (Low/High rate of introductions)** | 0.91 (0.88 - 0.93) | 0.99 (0.97 - 0.99) | 0.98 (0.96 - 0.99) | 0.97 (0.96 - 0.99) | 0.94 (0.93 - 0.96) | 0.98 (0.97-0.99) | 0.99 (0.97 - 0.99) |
|  | 0.65 (0.60 - 0.69) | 0.83 (0.79 - 0.86) | 0.81 ( 0.77 - 0.84) | 0.82 (0.79 - 0.85) | 0.76 (0.72 - 0.80) | 0.79 (0.75 - 0.82) | 0.88 (0.85 - 0.91) |
| **Reduced time from reactive vaccination to infection (Low/High rate of introductions)** | 0.91 (0.88 - 0.94) | 0.97 (0.96 - 0.99) | 0.96 (0.94 - 0.97) | 0.95 (0.93 - 0.97) | 0.90 (0.87 - 0.93) | 0.95 ( 0.93 - 0.97) | 0.99 (0.97 - 1.00) |
|  | 0.66 (0.62 - 0.70) | 0.85 (0.82 - 0.88) | 0.83 (0.79 - 0.86) | 0.79 (0.75 - 0.82) | 0.78 (0.74 -0.81) | 0.76 (0.72 - 0.80)) | 0.82 (0.78 - 0.85) |
| **Lower Vaccination coverage (Low/High rate of introductions)** | 0.91 (0.88 - 0.93) | 0.97 (0.95 - 0.99) | 0.94 (0.92 - 0.96) | 0.95 (0.92 - 0.97) | 0.94 (0.92 - 0.96) | 0.97 (0.95 - 0.98) | 0.97 (0.95 - 0.98) |
|  | 0.68 (0.64 - 0.72) | 0.81 (0.77 - 0.85) | 0.81 (0.77 - 0.85) | 0.77 (0.72 - 0.80) | 0.74 (0.70 - 0.78) | 0.76 (0.72 - 0.79) | 0.81 (0.77 - 0.84) |
| **Higher Vaccination coverage (Low/High rate of introductions)** | 0.91 (0.89 - 0.94) | 0.99 (0.98 - 1.00) | 0.99 (0.97 - 1.00) | 0.97 (0.95 - 0.98) | 0.92 (0.90 - 0.95) | 1.00 (0.99 - 1.00) | 0.99 (0.98 - 1.00) |
|  | 0.68 (0.66 - 0.71) | 0.88 (0.85 - 0.91) | 0.90 (0.87 - 0.93) | 0.88 (0.85 - 0.91) | 0.81 (0.77 - 0.84) | 0.78 (0.74 - 0.81) | 0.92 (0.90 - 0.95) |
| **Later date of intervention**<br>**(Low/High rate of introductions)** | 0.88 (0.85 - 0.91) | 0.98 (0.96 - 0.99) | 0.94 (0.92 - 0.96) | 0.91 (0.88 - 0.93) | 0.87 (0.84 - 0.90) | 0.93 (0.90 - 0.95) | 0.92 (0.90 - 0.95) |
|  | 0.64 (0.59 - 0.68) | 0.82 (0.79 - 0.86) | 0.81 (0.77 - 0.84) | 0.77 (0.74 - 0.81) | 0.76 (0.72 - 0.79) | 0.70 (0.66 - 0.74) | 0.84 (0.80 - 0.87) |

**Table 4:**
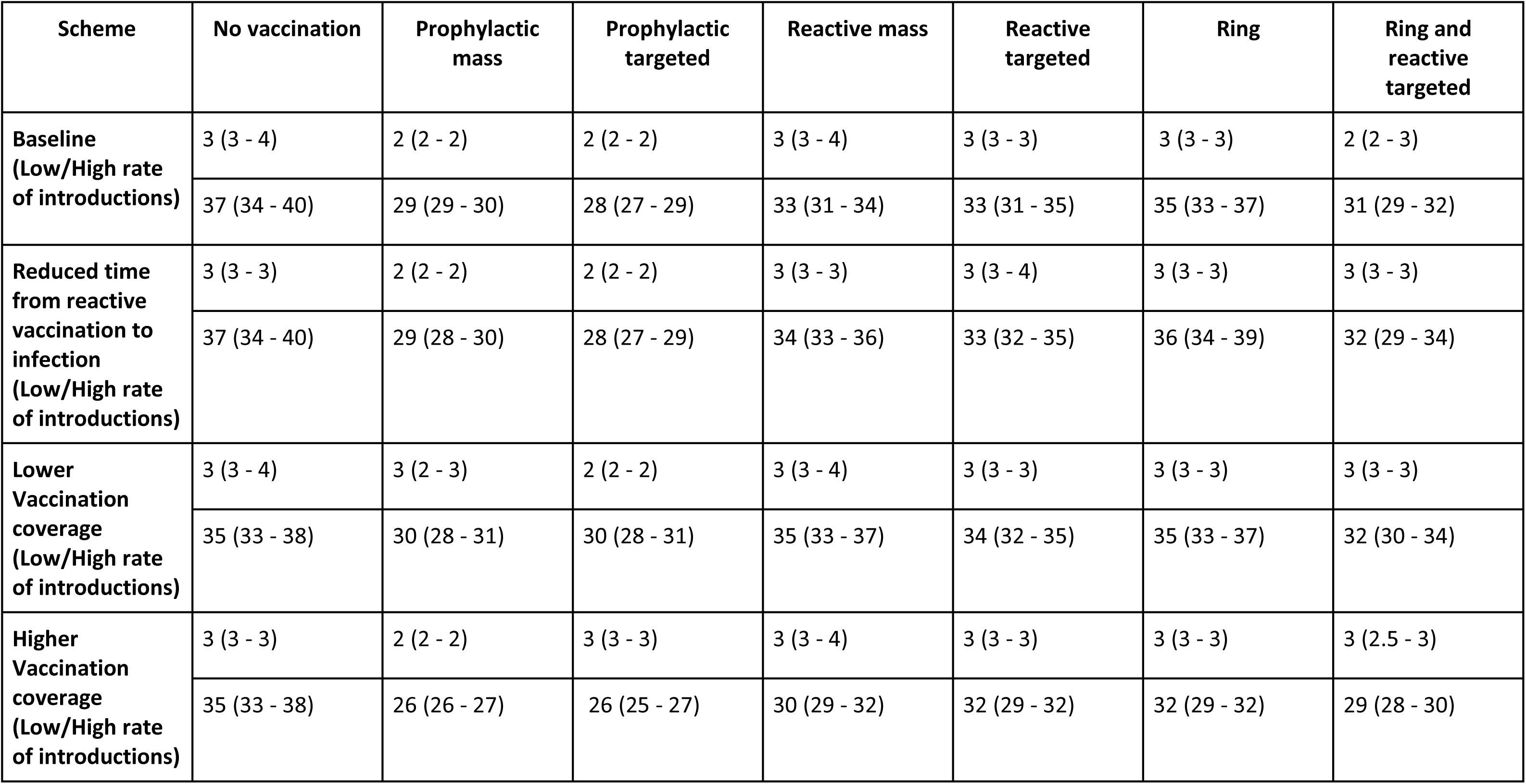
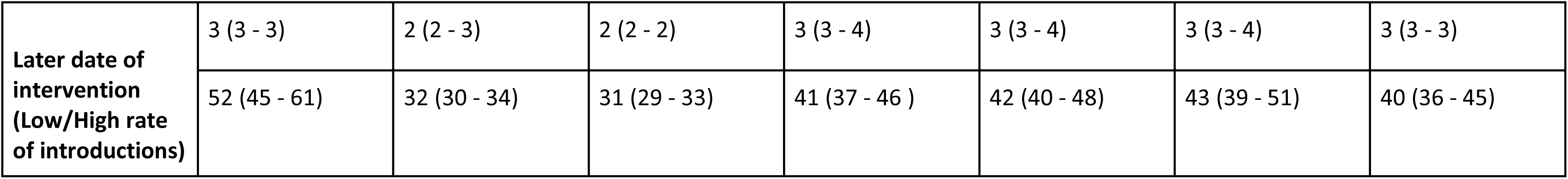
Median number of MVD cases predicted during different vaccination schemes

| **Scheme** | **No vaccination** | **Prophylactic mass** | **Prophylactic targeted** | **Reactive mass** | **Reactive targeted** | **Ring** | **Ring and reactive targeted** |
| --- | --- | --- | --- | --- | --- | --- | --- |
| **Baseline**<br>**(Low/High rate of introductions)** | 3 (3 - 4) | 2 (2 - 2) | 2 (2 - 2) | 3 (3 - 4) | 3 (3 - 3) | 3 (3 - 3) | 2 (2 - 3) |
|  | 37 (34 - 40) | 29 (29 - 30) | 28 (27 - 29) | 33 (31 - 34) | 33 (31 - 35) | 35 (33 - 37) | 31 (29 - 32) |
| **Reduced time from reactive vaccination to infection**<br>**(Low/High rate of introductions)** | 3 (3 - 3) | 2 (2 - 2) | 2 (2 - 2) | 3 (3 - 3) | 3 (3 - 4) | 3 (3 - 3) | 3 (3 - 3) |
|  | 37 (34 - 40) | 29 (28 - 30) | 28 (27 - 29) | 34 (33 - 36) | 33 (32 - 35) | 36 (34 - 39) | 32 (29 - 34) |
| **Lower Vaccination coverage**<br>**(Low/High rate of introductions)** | 3 (3 - 4) | 3 (2 - 3) | 2 (2 - 2) | 3 (3 - 4) | 3 (3 - 3) | 3 (3 - 3) | 3 (3 - 3) |
|  | 35 (33 - 38) | 30 (28 - 31) | 30 (28 - 31) | 35 (33 - 37) | 34 (32 - 35) | 35 (33 - 37) | 32 (30 - 34) |
| **Higher Vaccination coverage**<br>**(Low/High rate of introductions)** | 3 (3 - 3) | 2 (2 - 2) | 3 (3 - 3) | 3 (3 - 4) | 3 (3 - 3) | 3 (3 - 3) | 3 (2.5 - 3) |
|  | 35 (33 - 38) | 26 (26 - 27) | 26 (25 - 27) | 30 (29 - 32) | 32 (29 - 32) | 32 (29 - 32) | 29 (28 - 30) |
| Later date of intervention<br>(Low/High rate of introductions) | 3 (3 - 3) | 2 (2 - 3) | 2 (2 - 2) | 3 (3 - 4) | 3 (3 - 4) | 3 (3 - 4) | 3 (3 - 3) |
|  | 52 (45 - 61) | 32 (30 - 34) | 31 (29 - 33) | 41 (37 - 46 ) | 42 (40 - 48) | 43 (39 - 51) | 40 (36 - 45) |

After simulating a scenario with reduced coverage (20% less than baseline values) we found that, while no significant change in the median number of cases was observed, the proportion of controlled outbreaks decreased in the combined reactive and ring vaccination approach (Tables 3 and 4). This decrease was only apparent if the rate of introductions was high. Other vaccination approaches were not impacted by reducing vaccine coverage.

With increased coverage (20% greater than baseline values), all vaccination strategies performed significantly better than the no vaccination control, with both prophylactic approaches, as well as the combination of reactive targeted and ring vaccination performing best (Tables 3 and 4).

After simulating a scheme where the date of intervention was increased to 90 days after onset of the first case, there was a decrease in proportion of controlled outbreaks for reactive vaccination strategies at the higher introduction rate (Table 3). For instance, whereas 99% of outbreaks on average were controlled under a combined ring and reactive targeted vaccination scheme when interventions occurred after 21 days, this decreased to 84% with a delay of 90 days. This also translates to an increase in the median number of cases for these reactive vaccination approaches. Again taking the combined ring and reactive approach, we observed a small but significant increase from 31 (baseline) to 40 cases.

## Discussion

We combined and analysed data from each of the known MVD outbreaks to characterise key epidemiological parameters and assess the potential impact of a range of vaccination strategies. We found that the reproduction number of MVD in human populations is often low but very variable, consistent with the small and mostly self-limited nature of most outbreaks. Across all outbreaks, our estimates of *R*_0_ are lower than that calculated by Ajelli and Merler (25) (1.59; 95% CI: 1.53-1.66). This is largely because their estimates were based on data from the peak of the Angola outbreak alone – the largest recorded outbreak – whereas ours include all of the available data. Our estimated serial interval (9.2 days with a standard deviation of 4.4 days) is comparable to the generation time estimated by Ajelli and Merler from non-human primates (25) and shorter than that of the Ebola Virus disease (Zaire species) – see Table 1.

Although our estimates of the reproduction number after interventions are generally low, this is no guarantee that all future outbreaks will be limited in size. Our estimate of the dispersion parameter for *R0* (0.52 - 0.67) indicates that there is the potential for superspreading events to occur. Large future outbreaks of MVD cannot be ruled out. As the Angolan outbreak showed, for instance, a large outbreak can arise from a very limited number of introductions.

There are likely many social, epidemiological and environmental factors that may influence outbreak size. For instance, the two largest MVD outbreaks recorded to date (DRC in 1998-2000 and Angola in 2004-2005) occurred in populations that had recently been affected by civil war (16,18,20), which likely resulted in fragile health systems unable to prevent or rapidly control outbreaks.

To help understand what role vaccines might play in controlling future outbreaks we developed a simple branching process model and parameterised it from our analyses of the epidemiology. As expected, vaccination generally increased the probability of outbreaks being controlled compared to no

vaccination. Over the range of strategies and parameter values considered, generally similar effects were achieved. Exceptions included reactive targeted vaccination when the rate of spill-over introductions is low, as well as mass vaccination strategies when only very low coverage (e.g. 30%) is achieved.

Vaccination could also be expected to reduce the median outbreak size, though this reduction is often relatively small since the median number of cases for the no vaccination scheme is already low (3 when the rate of introduction is low, 37 when high).

A combined ring and targeted vaccination was generally the most effective option since it results in a high probability of control and a low median outbreak size. However, if there are few introductions, the added effect of targeted vaccination over ring vaccination alone is negligible. Nevertheless, the combined approach might still be preferred since the rate of spill-over introductions might be difficult to assess in real-time without comprehensive sequence data.

Quantifying the number of hypothetical vaccines that may have been required to help control the previous 15 outbreaks of MVD is difficult as the target population is unclear for most of the outbreaks. However, it is possible to obtain a rough idea of the number of doses that may be required for the different strategies for a few of the outbreaks. The DRC outbreak of 1998-2000 only affected two towns with a combined population of approximately 85,000 (28). Had a vaccine been available the number of vaccines required for a targeted strategy was 24,000, assuming 40% of the population were miners (i.e. 80% of males) and a 70% coverage rate. A mass vaccination strategy with 50% coverage, on the other hand, would require 43,000 vaccines. This outbreak does, however, constitute an extreme case where a large proportion of the population were considered high-risk individuals and thus candidates for receiving a vaccine under a targeted vaccination scheme. In comparison, the outbreak in Uganda, 2012, spanned 3 of the country’s districts and was instigated by only a single known zoonotic case. Although it is not known what fraction of the population was high risk, applying the fraction of miners and healthcare workers in the country as a whole to these districts gives a possible indication (i.e. 5,600 high risk individuals amongst a total population of 730,000) (12). Hence, 3,900 vaccines would have been required under a targeted vaccination scheme with 70% coverage and 370,000 vaccines (two orders of magnitude greater) would have been required under a mass vaccination scheme with 50% coverage.

Although efforts are ongoing to develop MVD vaccines (4), the results of our study suggest that it may be difficult to carry out Phase 3 trials, since we predict that few cases will be observed in a typical outbreak, and these may well be rapidly controlled by other interventions. To counter this problem, the World Health Organisation has developed a Core Protocol approach that is designed to allow trial results to be combined across multiple outbreaks to accrue sufficient data and statistical power to assess vaccine effectiveness (26, 29). There has also been a recent paper from WHO on a core protocol for estimating Marburg virus vaccine efficacy (29). The results of our model could be helpful in estimating how many cases and outbreaks may be necessary to include in such a longitudinal multi-outbreak study.

A major limitation of our study is the lack of data due to the infrequency of MVD outbreaks, with most being relatively small, and in many cases, scant availability of epidemiologic data. This paucity of data and heterogeneity across and within outbreaks leads to wider credible intervals associated with *R_0_* and *E*. Another limitation is that, while we calculated and used a constant rate of zoonotic introductions, in reality, the rate often varies as a function of time. For example, the outbreak in the DRC appears to have been driven by seasonal introductions into miners (6). However, due to a lack of data on zoonotic introductions into specific persons, we opted for model simplicity and chose a constant rate for each outbreak.

We chose to use a simple branching process model that does not include, for instance, depletion of the susceptible population. Depletion of susceptibles may well be an important consideration, especially under a mass vaccination strategy. Nor did we account for any possible waning of immunity post-vaccination. At present there is a lack of data on the effectiveness of vaccination - we have informed this part of our analysis by making broad and simplistic assumptions. Several vaccines are currently in the pipeline (4) and as results emerge we can update our work accordingly. Also, our study uses a branching process model that forces one case of MVD at time *t* = *0*. However, both prophylactic vaccination approaches may, indeed, prevent many outbreaks from even beginning. These are not considered in our model.

Our study shows that various vaccination strategies can be effective in helping to control outbreaks of MVD, with the best approach varying with the particular epidemiologic circumstances of each outbreak. Of course, many logistical and economic factors must be considered. Further studies on the economic factors involved in vaccinating against MVD will be required but are beyond the scope of this study. Given the rarity and generally small size of MVD outbreaks, prophylactic mass vaccination of large populations is unlikely to be feasible or warranted. However, as has been proposed for vaccination for Ebola virus, vaccination for relatively infrequent but dangerous emerging infectious diseases might be incorporated into comprehensive vaccination for numerous diseases, serving as a driver of broader health systems strengthening (30). The rationale for this approach would be further strengthened by development of pan-filovirus vaccines, for which research is underway (31), especially if protection is long-lasting.

## Supporting information

Technical Appendix

## Data Availability

All data produced in the present study are available upon reasonable request to the authors.

## Ethical Approval

Ethical approval for the study was granted by the London School of Hygiene & Tropical Medicine Ethics Committee (Reference Number 26566).

## Acknowledgements

This research is funded by the Department of Health and Social Care using UK Aid funding and is managed by the National Institute for Health and Care Research (VEEPED: PR-OD-1017-20002). The views expressed in this publication are those of the authors and not necessarily those of the Department of Health and Social Care.

## Supporting Information

Technical Appendix: Supporting Information to the paper

