## Supplementary material for "Modelling Vaccination Strategies for the Control of Marburg Virus Disease Outbreaks": Technical Appendix

#### Branching Process Transmission Model

We created a branching process model to simulate the transmission of marburgvirus disease (MVD), following a similar method to that in (1). Secondary cases were generated from a Poisson distribution, whose mean was the force of infection calculated at any given point in time. Hence, given an epidemiological curve at timestep $t$, the number of cases at $(t + 1)$is:

$$y_t \sim Poisson(\lambda_t)$$

with

$$\lambda_t = R\, \Sigma_{s \leq t}\, y_s\, w(t - s)$$

where $R$ is the reproduction number, and $w(.)$ the probability mass function from the discretised serial interval distribution.

The rate of zoonotic introductions were also Poisson distributed, with mean equal to the rate of introduction observed in data from previous MVD outbreaks. Each case was also categorised as being reported or unreported; reported cases reduced the basic reproduction number, $R_{basic}$ , by the *intervention efficacy*, IE, a number that incorporates the effects of contact tracing and case isolation. Thus:

$$R_{Unreported} = R_{basic}$$
$$R_{Reported} = R_{basic}(1 - IE)$$

Using an Approximate Bayesian Computation (ABC) approach, we obtained the posterior distribution of [$R_{basic}$, IE] pairs for each previous outbreak. 100 of these pooled posteriors were randomly drawn and used to simulate 3 vaccination strategies: ring, targeted and mass vaccination, as well as a combination of these.

#### Vaccine Efficacy

Vaccine efficacy (VE) was modelled as a function of the elapsed time ($t$) between the vaccination and infection of any case. Specifically, we chose a logistic model of the form:

$$VE(t) = \frac{\phi_1}{1 + exp(\frac{\phi_2 - t}{\phi_3})}$$

where $\phi_1$ is the maximum vaccine efficacy, $\phi_2$ the time at which the inflection point of the logistic curve occurs, and $\phi_3$ the scaling parameter.

From recent results of the VSV-MARV Marburg vaccine (2), the maximal VE, $VE_{max} \approx 1$ in NHPs. From the same study, the time from vaccination to $VE_{max}$ was 7 days. We used a nonlinear least squares approach to fit the 2 piecewise functions (the increase in VE from 0 to $VE_{max}$ and the constant VE once this had been reached).

Hence, VE comprises 3 parts: vaccination delay (where $VE = 0$), a period when VE gradually increases and, finally, a period of maximal protection. To simplify our model, we assumed VE to be the mean of these values across the duration of a cluster of MVD cases (which was 30 days, according to data from the DRC outbreak).

For prophylactic vaccination strategies, samples of VE are taken from the upper portion of this logistic curve: specifically, we assumed an average of 20 days (s.d. 5 days)
For reactive vaccination strategies, samples of VE are taken from a wider range of the logistic curve (mainly its slope). Specifically, we assumed an average of 9 days (s.d. 4 days) between vaccination and infection.

The reproduction number for individuals who had been vaccinated, $R_{Vaccinated}$, (bearing in mind that these people must also have benefitted from other interventions), was:

$$R_{Vaccinated} = R_{basic}(1 - IE)(1 - VE)$$

where: $R_{basic}$ is the basic reproduction number (before interventions and vaccinations have taken place) and $IE$ the intervention efficacy.

#### Estimates of the dispersion parameter, $k$

18 chains of transmission were identified in the DRC outbreak. Broadly speaking, we estimated the dispersion parameter by fitting a negative binomial distribution to the lengths of these chains. More specifically, we obtained a range of estimates for $k$, depending on whether the index case of a chain was a miner. If not, we supposed there was a 67% probability that the index case had been missed; the figure reported in (4). There were also missed cases in 2 identified chains which only had length 1, where we could be certain that there were incomplete data. Missed cases implied lengthening the chain by 1. We, thus, estimated the dispersion parameter as: $0.52 < k < 0.67$.

#### Serial interval

The serial interval can be modelled as a gamma distribution with a mean of 9.2 days and standard deviation of 4.4 days. Figure A2 shows a plot of observed and fitted values.

### Figures

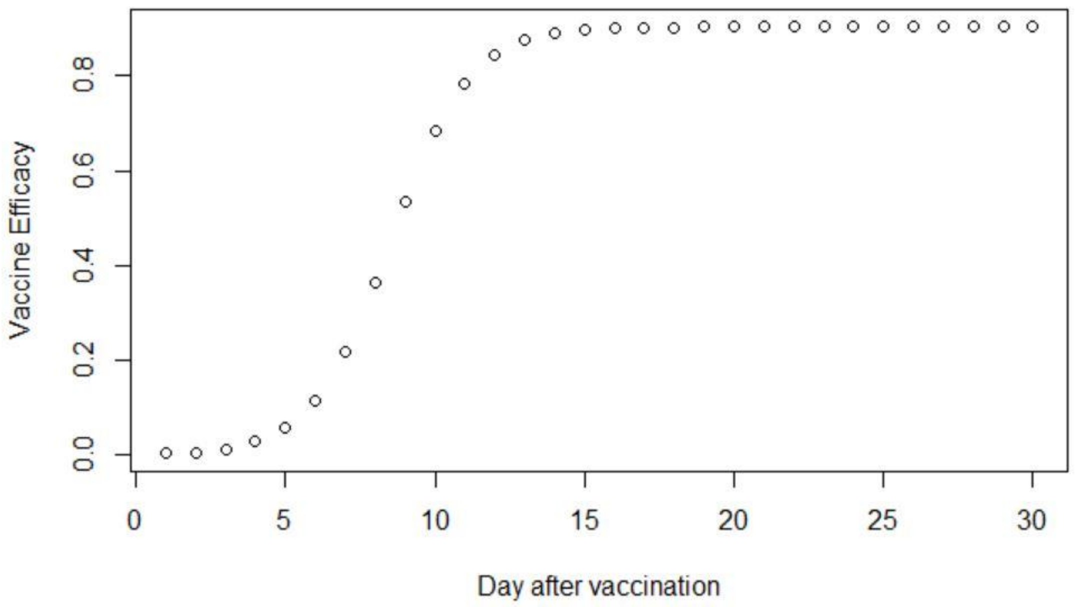


**Figure A1:** Curve showing vaccine efficacy as a logistic function of days after vaccination

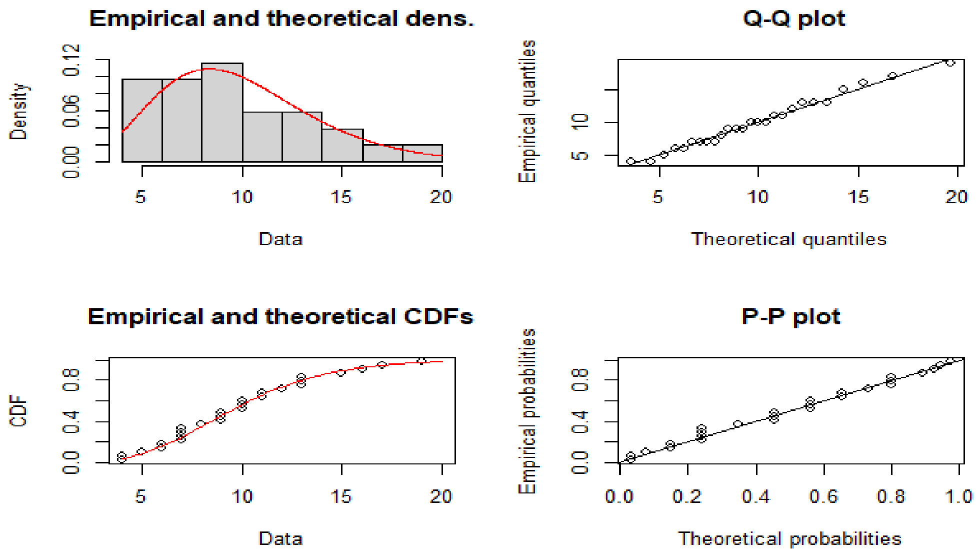


**Figure A2:** Serial interval, modelled as a gamma distribution

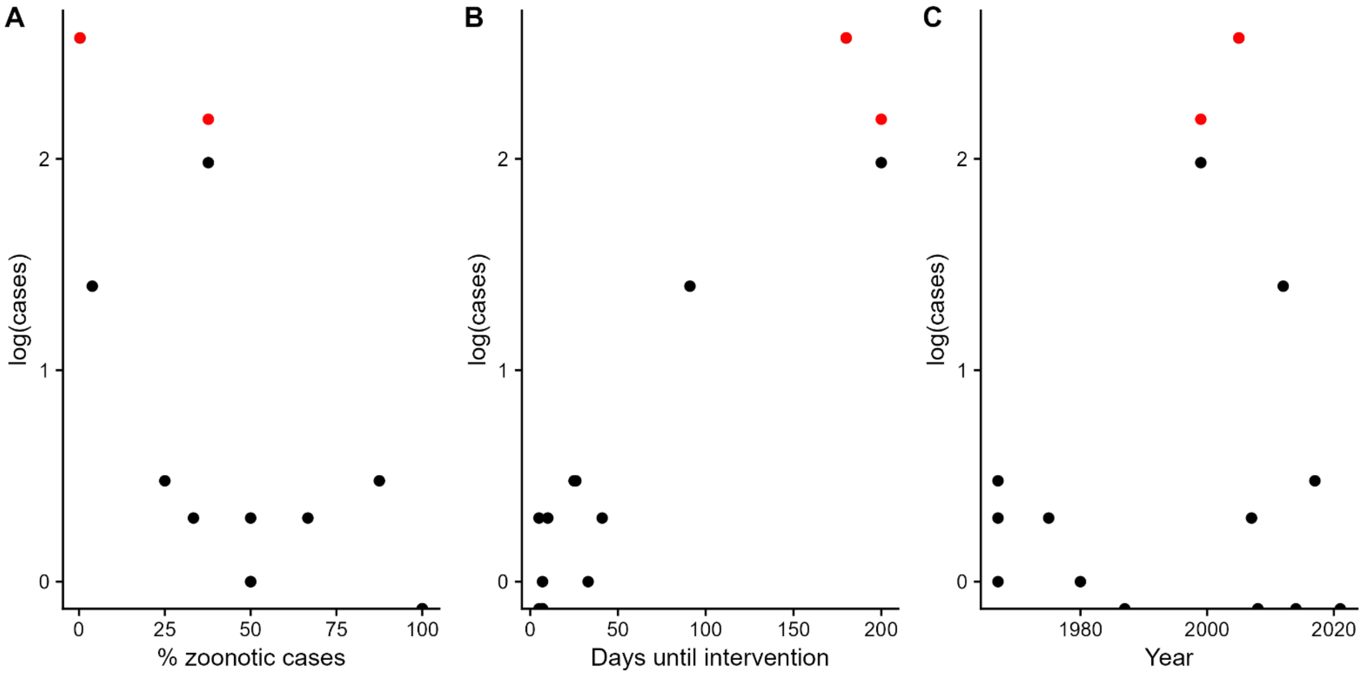


**Figure A3:** Scatterplots showing the logarithm (base 10) of the number of secondary marburgvirus cases (confirmed and probable) according to: A. proportion of zoonotic cases, B. days until intervention and C. year of outbreak. Red dots indicate outbreaks affected by civil war (namely, DRC and Angola).

### Tables

**Table A1:** Previous Marburgvirus outbreaks

| Outbreak Location | Year(s) | Probable and Confirmed Cases | Deaths |
|---|---|---|---|
| **Marburg, Germany** | 1967 | 24 | 5 |
| **Frankfurt, Germany** | 1967 | 6 | 2 |
| **Belgrade, Yugoslavia** | 1967 | 2 | 0 |
| **Johannesburg, South Africa** | 1975 | 3 | 1 |
| **Nairobi, Kenya** | 1980 | 2 | 1 |
| **Nairobi, Kenya** | 1987 | 1 | 1 |
| **Durba and Watsa, Democratic Republic of Congo** | 1998-2000 | 154 | 128 |
| **Uige, Angola** | 2004-2005 | 374 | 329 |
| **Uganda** | 2007 | 4 | 2 |
| **USA (via Uganda)** | 2008 | 1 | 0 |
| **Netherlands (via Uganda)** | 2008 | 1 | 1 |
| **Uganda** | 2012 | 26 | 15 |
| **Uganda** | 2014 | 1 | 1 |
| **Uganda** | 2017 | 4 | 1 |
| **Guinea** | 2021 | 1 | 1 |

**Table A2:** Median and (in parentheses) 95% credible intervals of the basic reproduction number ($R_0$) and intervention efficacy ($E$) for each MVD outbreak

| Outbreak Location | Year(s) | Basic reproduction number, $R_0$ | Intervention Efficacy, ($E$) | Reproduction number after intervention |
|---|---|---|---|---|
| **Marburg, Germany** | 1967 | 0.51 (0.049 – 1.8) | 0.64 (0.035 – 0.98) | 0.17 (0.006 – 0.70) |
| **Frankfurt, Germany** | 1967 | 0.86 (0.092 - 2.6) | 0.50 (0.023 - 0.97) | 0.36 (0.014 – 1.8) |
| **Belgrade, Yugoslavia** | 1967 | 0.89 (0.050 - 2.8) | 0.61 (0.036 - 0.98) | 0.27 (0.007 – 1.6) |
| **Johannesburg, South Africa** | 1975 | 0.91 (0.049 – 2.7) | 0.59 (0.032 – 0.97) | 0.29 (0.008 – 1.7) |
| **Nairobi, Kenya** | 1980 | 0.62 (0.019 – 2.1) | 0.47 (0.034 – 0.96) | 0.25 (0.005 – 1.6) |
| **Nairobi, Kenya** | 1987 | 0.68 (0.027 – 2.6) | 0.51 (0.024 – 0.98) | 0.27 (0.003 – 1.8) |
| **Durba and Watsa, Democratic Republic of Congo** | 1998-2000 | 0.76 (0.57-0.98) | 0.44 (0.024 - 0.97) | 0.42 (0.022 – 0.72) |
| **Uige, Angola** | 2004-2005 | 1.2 (1.0 – 1.9) | 0.54 (0.036 - 0.97) | 0.58 (0.031 – 1.5) |
| **Uganda** | 2007 | 0.58 (0.037 – 2.3) | 0.52 (0.045 – 0.98) | 0.22 (0.006 – 1.3) |
| **USA (via Uganda)** | 2008 | 0.66 (0.021 – 2.6) | 0.49 (0.022 – 0.97) | 0.27 (0.005 - 1.7) |
| **Netherlands (via Uganda)** | 2008 | 0.69 (0.031 – 2.6) | 0.50 (0.039 – 0.97) | 0.29 (0.006 - 1.7) |
| **Uganda** | 2012 | 1.1 (0.66 – 1.9) | 0.55 (0.023 – 0.97) | 0.48 (0.029 – 1.4) |
| **Uganda** | 2014 | 0.68 (0.027 – 2.6) | 0.51 (0.024 – 0.98) | 0.26 (0.003 – 1.8) |
| **Uganda** | 2017 | 0.67 (0.020 – 2.5) | 0.53 (0.027 – 0.97) | 0.27 (0.003 – 1.6) |

| | | | | |
|---|---|---|---|---|
| **Guinea** | 2021 | 0.84 (0.036 – 2.8) | 0.49 (0.016 – 0.98) | 0.006 (0.32 – 1.9) |
